# SARS-CoV-2 under an elimination strategy in Hong Kong

**DOI:** 10.1101/2021.06.19.21259169

**Authors:** Haogao Gu, Ruopeng Xie, Dillon C. Adam, Joseph L.-H. Tsui, Daniel K. Chu, Lydia D.J. Chang, Sammi S.Y. Cheuk, Shreya Gurung, Pavithra Krishnan, Daisy Y.M. Ng, Gigi Y.Z. Liu, Carrie K.C. Wan, Kimberly M. Edwards, Kathy S.M. Leung, Joseph T. Wu, Dominic N.C. Tsang, Gabriel M. Leung, Benjamin J. Cowling, Malik Peiris, Tommy T.Y. Lam, Vijaykrishna Dhanasekaran, Leo L.M. Poon

**Affiliations:** School of Public Health, LKS Faculty of Medicine, The University of Hong Kong, Hong Kong, China; HKU-Pasteur Research Pole, School of Public Health, LKS Faculty of Medicine, The University of Hong Kong, Hong Kong, China; Laboratory of Data Discovery for Health, Hong Kong Science and Technology Park, Hong Kong, China; Centre for Health Protection, Department of Health, The Government of Hong Kong Special Administrative Region, Hong Kong, China; Centre for Immunology & Infection, Hong Kong Science and Technology Park, Hong Kong, China

## Abstract

Hong Kong utilized an elimination strategy with intermittent use of public health and social measures and increasingly stringent travel regulations to control SARS-CoV-2 transmission. By analyzing >1700 genome sequences representing 17% of confirmed cases from 23-January-2020 to 26-January-2021, we reveal the effects of fluctuating control measures on the evolution and epidemiology of SARS-CoV-2 lineages in Hong Kong. Despite numerous importations, only three introductions were responsible for 90% of locally-acquired cases, two of which circulated cryptically for weeks while less stringent measures were in place. We found that SARS-CoV-2 within-host diversity was most similar among transmission pairs and epidemiological clusters due to a strong transmission bottleneck through which similar genetic background generates similar within-host diversity.

**One sentence summary:** Out of the 170 detected introductions of SARS-CoV-2 in Hong Kong during 2020, three introductions caused 90% of community cases.

---

Severe acute respiratory coronavirus 2 (SARS-CoV-2) emerged in late 2019 (*1*) and has caused over 170 million confirmed cases and over three million deaths worldwide (as of 1-July-2021) (*2*). Heterogeneity in disease severity (*3-5*) and high virus transmission rates (*6, 7*) necessitated extensive and diverse control strategies, which achieved varied degrees of success. Hong Kong (population 7.5 million) is among the few regions that have been relatively successful in prevention and control of community transmission using an elimination strategy (*8*), with intermittent public health and social measures combined with strict isolation of cases and quarantine of close contacts in designed facilities (*9, 10*). Increasingly stringent travel regulations have been implemented to limit importation of infections (**Fig. 1**). As of 1-July-2021, 11,855 laboratory confirmed cases have been detected resulting in 210 deaths. Using contact tracing data from January to April 2020 (waves one and two) we have shown that sustained community transmission in Hong Kong was largely driven by superspreading events, with 80% of community infection caused by 20% of cases (*10*). However, two large outbreaks have occurred in Hong Kong since this period (waves three and four). Here, we show that the three major surges in infections that occurred during waves two to four were the result of only three virus introductions (PANGO lineages (*11*) B.3, B.1.1.63, and B.1.36.27) out of a total of 170 captured through genome sequencing, resulting in 90.0% of the confirmed cases of SARS-CoV-2 observed in the community in Hong Kong. Using genomic data generated from travel-related (n=186) and community cases (n=1,713) during all four waves of the pandemic, comprising 51.4%, 21.1%, 23.6% and 13.7% of all cases in the corresponding period, respectively (**figs. S1, S2** and **S3, tables S1** and **S2**), we discuss the effects of intermittent use of public health and social measures on the evolution and epidemiology of SARS-CoV-2 in Hong Kong.

## Introductions, local spread, and delayed detection during waves one and two

During wave one (23-January-2020 to 22-February-2020), laboratory-confirmed cases did not exceed 10 per day (**Fig. 1** and **table S1**) and were comprised of both travel-related (n=17/70, 24.3%) and community-acquired cases (n=53/70, 75.7%). The first recognised introduction was detected on 30-January-2020 among a family cluster where the index case had returned from Wuhan, China on 22-January, while the first community case without a known source (travel history or contact with a confirmed case) was also reported on 30-January. However, among two of the earliest locally circulating lineages, time-scaled phylogenetic analysis estimated a median time to most recent common ancestor (tMRCA) of 30-December-2019 (95% Highest Posterior Density Interval (HPD) 24-December-2019 – 1-January-2020) indicating direct ancestry to cases circulating prior to the earliest recognised introductions (**Fig. 2A**). Critically, these lineages had no recognised epidemiological or phylogenetic link to all other imported cases identified and sampled at the time, indicating that the source of these lineages initially entered Hong Kong undetected and triggered sustained community transmission during the first wave. Though the tMRCA indicates introduction could have occurred as early as 24-December-2019, which would represent one of earliest examples of transmission outside mainland China, it cannot be proven as the lineage diversity observed may have first accumulated in mainland China and subsequently been imported closer to the detection of the earliest cases.

**Fig. 1.**
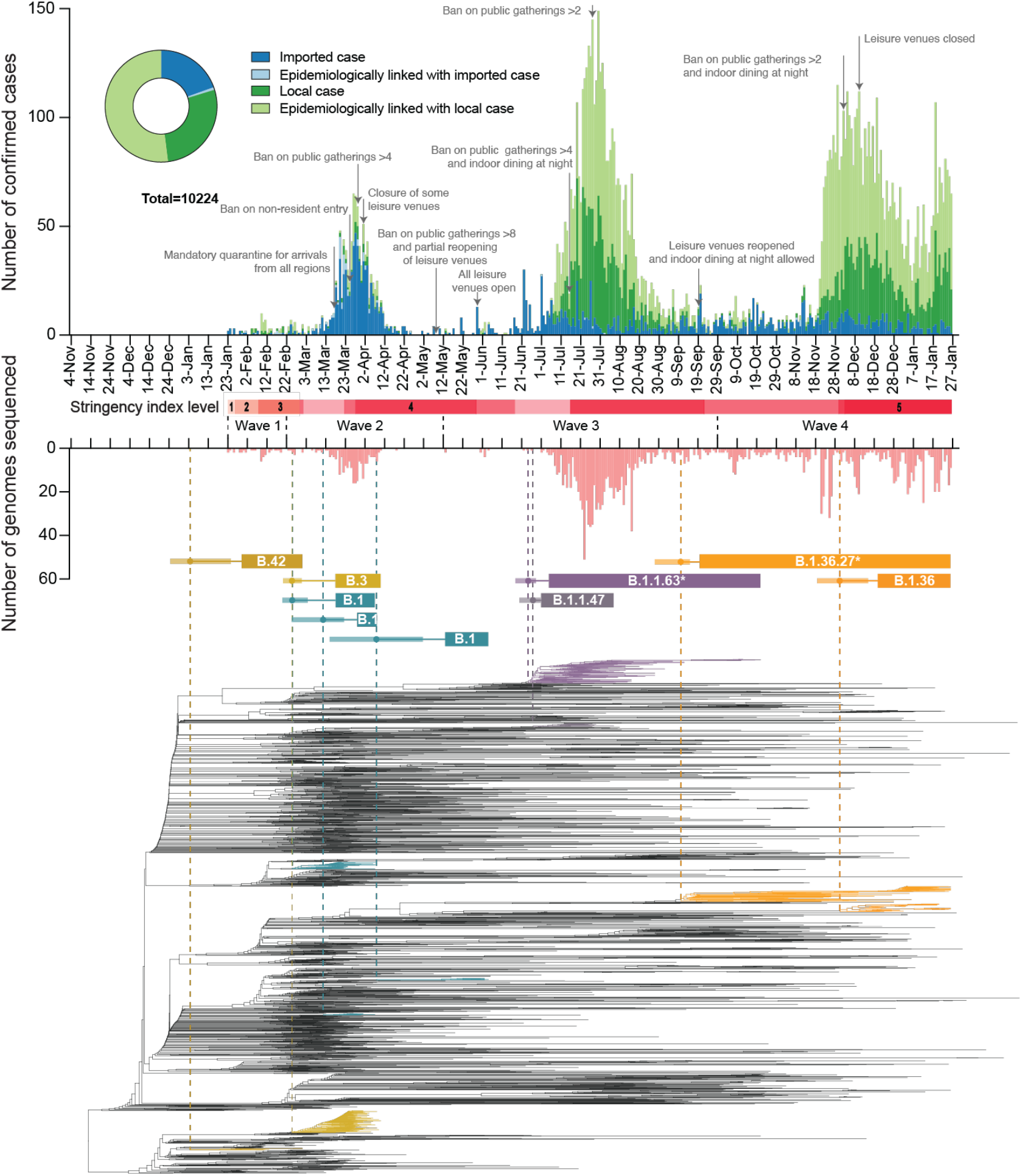
Epidemiological summary and time-scaled phylogeny of SARS-CoV-2 in Hong Kong. The number of genomes sequenced in this study is shown below in light red. Time-scaled phylogeny of SARS-CoV-2 genomes from Hong Kong, in which lineages HK-wave3 and HK-wave4A were subsampled to 100 and 65 sequences, showing B.1.1.63* and B.1.36.27*, respectively. *denotes the real clade contains more than one PANGO lineage (see **Materials and Methods** and **table S4**). Red shaded bars below the timeline delineate five stringency levels of control measures in Hong Kong based on the Oxford COVID-19 Government response tracker (level 1: <40; level 2 : 40–50; level 3: 50–60; level 4: 60–70; level 5: >70) (*12*).

**Fig. 2.**
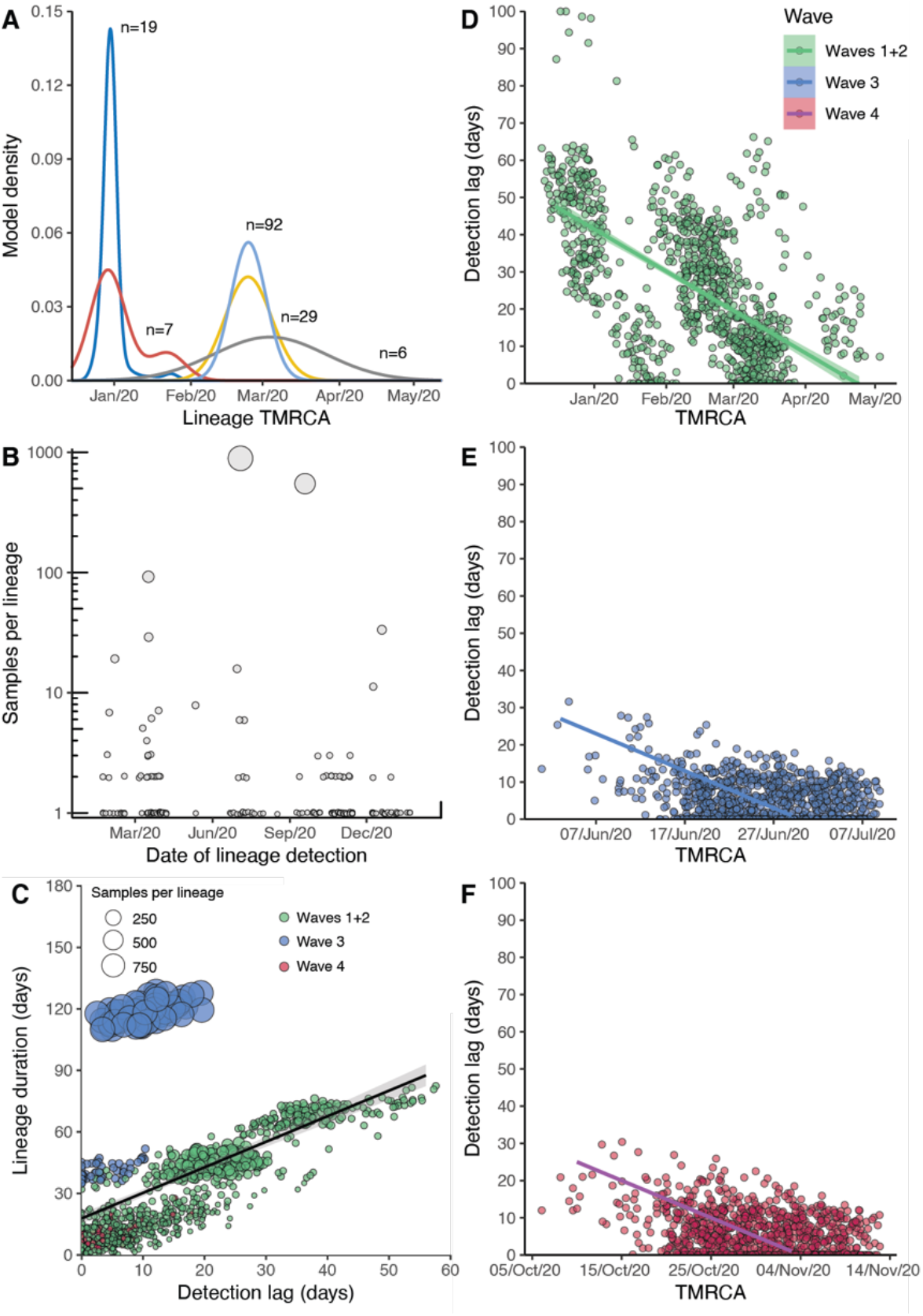
Descriptive and temporal dynamics of SARS-CoV-2 lineages in Hong Kong. **(A)** Time to most recent common ancestor (tMRCA) among the five earliest locally circulating lineages of SARS-CoV-2 introduced into Hong Kong. **(B)** Number of SARS-CoV-2 genomic samples per lineage identified over time. Lineage size is ordered on a log10 scale, and plotted by the earliest confirmation date among the lineages. **(C)** Correlation between the detection lag of locally circulating lineages and the final lineage duration. Points represent a random sample of 1,000 lineages from a Bayesian posterior tree distribution (n=8,000). **(D:E)** Detection lag over time as a function of tMRCA across three epidemic periods **(D)** waves one and two, **(E)** wave three, **(F)** wave four. Overall, a significant reduction in detection lag was observed over time and across each epidemic wave. Points in each panel represent a random sample of 1,000 lineages from a Bayesian posterior tree distribution (n=8,000).

As SARS-CoV-2 was declared a global pandemic on 11-March-2020, Hong Kong experienced a substantial rise in travel-related cases (n=705/978, 72.1%, **Fig. 1** and **table S1**) concomitant with large epidemics internationally (wave two). Introductions occurred mostly from outside of China (**fig. S4**), with a moderate increase observed in community cases (n=273/978, 27.9%). Again, the inferred common ancestry of community-linked lineages indicated circulation prior to or during early March 2020 (tMRCA), indicating a pattern of prolonged cryptic transmission that preceded increases in community spread (**Fig. 2A**). The largest of these local lineages comprised 92 genomes or 38.0% (n=92/242) of all sampled genomes during wave two and was classified as PANGO lineage B.3. This lineage was associated with a superspreading event from which contact tracing identified 106 community cases. Genomic analysis linked an additional 16 sporadic community cases, increasing the total inferred cluster size to 122 or 46.2% (n=122/326) of community cases during waves one and two.

Overall, during waves one and two, there were 38 lineages circulating in the community (out of a total of 61 unique imported clades). The median size for non-singleton local lineages was six cases, and the median duration of circulation was 10.5 days. Among local lineages without traced contact to an imported case, the median delay in lineage detection (time between lineage tMRCA and confirmation of the first detected case) was 25 days, though this was noticeably higher in January (median = 31 days), before significantly improving to eight days by early May 2020, likely related to early delays and subsequent improvements in case detection (Spearman’s test, rho (ρ) = -0.49, p < 0.001, **Fig. 2D**).

## Travel measures and the suppression of overseas introductions

Following a peak of infections during 16–27 March 2020 (wave two; **Fig. 1** and **table S1**), a rapid decline occurred as travel and community restrictions were expanded to include mandatory quarantine on arrival with strengthened testing, restrictions on non-resident entry, school closures, adjusted work arrangements, and bans on public gatherings (*13*). Travel restrictions began as early as 26-January-2020. First, all non-residents that visited Hubei province within two weeks were barred entry into Hong Kong. This was followed by a mandate for compulsory quarantine of passenger arrivals from regions affected with SARS-CoV-2, extending from mainland China to South Korea, Iran, Italy, and the Schengen region, and culminating in the barring of entry of non-residents during the peak of wave two in March. No community cases were reported from mid-April to 12-May-2020, and community measures were gradually relaxed to allow public gatherings of at most eight (from four) people with restricted opening of leisure venues (**Fig. 1**). Based on the Oxford COVID-19 Government response tracker (*12*), stringency of control measures reduced from level 4 to level 2 during this period (**Fig. 1**).

## Reintroductions and local surge during waves three and four

The first large SARS-CoV-2 outbreak in Hong Kong occurred from July to September 2020 (wave three), resulting in >4,000 cases (**table S1**). With a predominant number of cases in the community (n=3,385/4,032, 84.0%), this third epidemic wave was preceded by a period of increased detection of travel-related cases during July 2020 (similar to waves one and two, **Fig. 2B**). While importations during wave two were predominantly from Europe, subsequent cases were mostly from Asia (**fig. S4, table S3** and **data S1**). The number of laboratory-confirmed cases continued to rise, reaching a peak of >120 cases per day in late July (**Fig. 1**). Following the implementation of increasingly stringent public health and social distancing measures, the local epidemic subsided in September. However, beginning in early November a second resurgence (termed wave four) occurred, resulting in >6000 additional cases. Wave four peaked in December 2020 and slowly declined towards zero daily cases by April 2021.

Sequencing identified 170 virus lineages belonging to 71 PANGO lineages in Hong Kong within one year (**Fig. 2** and **data S2**). However, 87.0% of those lineages were detected only in travel-related cases or one community case, and no variants of concern were detected in the community during waves three and four. Notably, a single introduction belonging to lineage B.1.1.63 resulted in over 90.0% of genomes sampled during wave three (92.4%, 881/953), forming a HK-wave3 clade (n=902) (also see **fig. S1** and **table S4**). However, the earliest imported cases of the HK-wave3 clade were not sampled, indicating cryptic transmission prior to detection. In a similar trend, two lineages led to most of the fourth wave cases: over 74% genomes (552/704) formed one clade (HK-wave4A, B.1.36.27, **table S4**), and 4.7% (33/704) formed another (HK-wave4B, B.1.36).

By comparing the tMRCA of local non-singleton clusters in Hong Kong (n=7, 6.4% (7/109)) (**Fig. 2** and **data S3**), we identified two introductions (HK-wave3 and HK-wave4A) that circulated in the community for 108 and 128 days, respectively. The delay in detection of local non-singleton lineages remained low during waves three and four (mean=2.9 days, 95% HPD 0 - 13 days) (**Fig. 2E, F**), with a detection delay of 11 and 10 days for the two large clades, HK-wave3 and HK-wave4A, respectively (**data S3**). A negative correlation between delay over time from wave one to wave four (Spearman’s test, rho (ρ) = -0.72, p < 0.001), indicates improvements in identifying cases during 2020 (**Figs. 2D, E** and **F**). Interestingly, the cryptic circulation of HK-wave3 occurred during periods of reduced stringency (level 2), however HK-wave4A introduction occurred during the late stages of wave three when stringency level 4 was enacted, and continued to circulate cryptically when stringency level was reduced to level 2 on presumed suppression of the second wave. Similarly, HK-wave4B introduction occurred during relatively stringent level 4 (**Fig. 1**), while a significant positive correlation between increasing lag in lineage detection and lineage duration was observed (Spearman’s test, rho (ρ) = 0.70, p < 0.001, **Fig. 2C**).

## Contrasting patterns of epidemiology during waves three and four

To understand the effects of public health and social measures on community transmission, we characterized the two largest clades that circulated during waves three and four and identified contrasting pattens of genomic evolution and underlying transmission dynamics. HK-wave3 emerged during a period of reduced community measures (stringency level 2) and was controlled by increasing stringency to level 4. By contrast, HK-wave4A emerged under stringency level 4 and circulated cryptically during reduced level 3. Although the stringency level was rapidly raised to 5 on detection, HK-wave4A continued to circulate for several months.

The effective reproductive number (R_e_) estimated using a birth-death skyline serial (BDSS) model (*14*) for 902 HK-wave3 genomes collected from all 18 districts in Hong Kong (5-July-2020 to 21-October-2020) showed that R_e_ was significantly higher (∼3) from the tMRCA in late June or early July (mean tMRCA = 1-July-2020, 95% HPD, 26-June-2020 to 4-July-2020) until recognition of the lineage on 5-July when stringency level was 2 (**Fig. 3**), highlighting a period of rapid virus expansion when leisure venues were open and public gatherings of up to 50 people were allowed. Notably, as the control stringency was increased on 15-July (from level 2 to 4), R_e_ subsided to ∼2 and subsequently decreased below ∼1. An increasing number of cases continued to be reported among close contacts in residential care homes for the elderly or disabled, households, hospitals, dormitories and workplaces, whereas just 12.6% of the genomes were attributable to social interactions (**table S5**). Phylogenies reveal a rapid termination of transmission lineages among close contacts, leading to disappearance of all but one sub-lineage that continued to circulate with R_e_ >1 until extinction in October 2020. These results indicate suppression with level 4 stringency during wave three, complimented by aggressive contact tracing, resulted in the elimination of the majority of wave three transmission chains despite the intermittent rise in cases among close contacts.

**Fig. 3.**
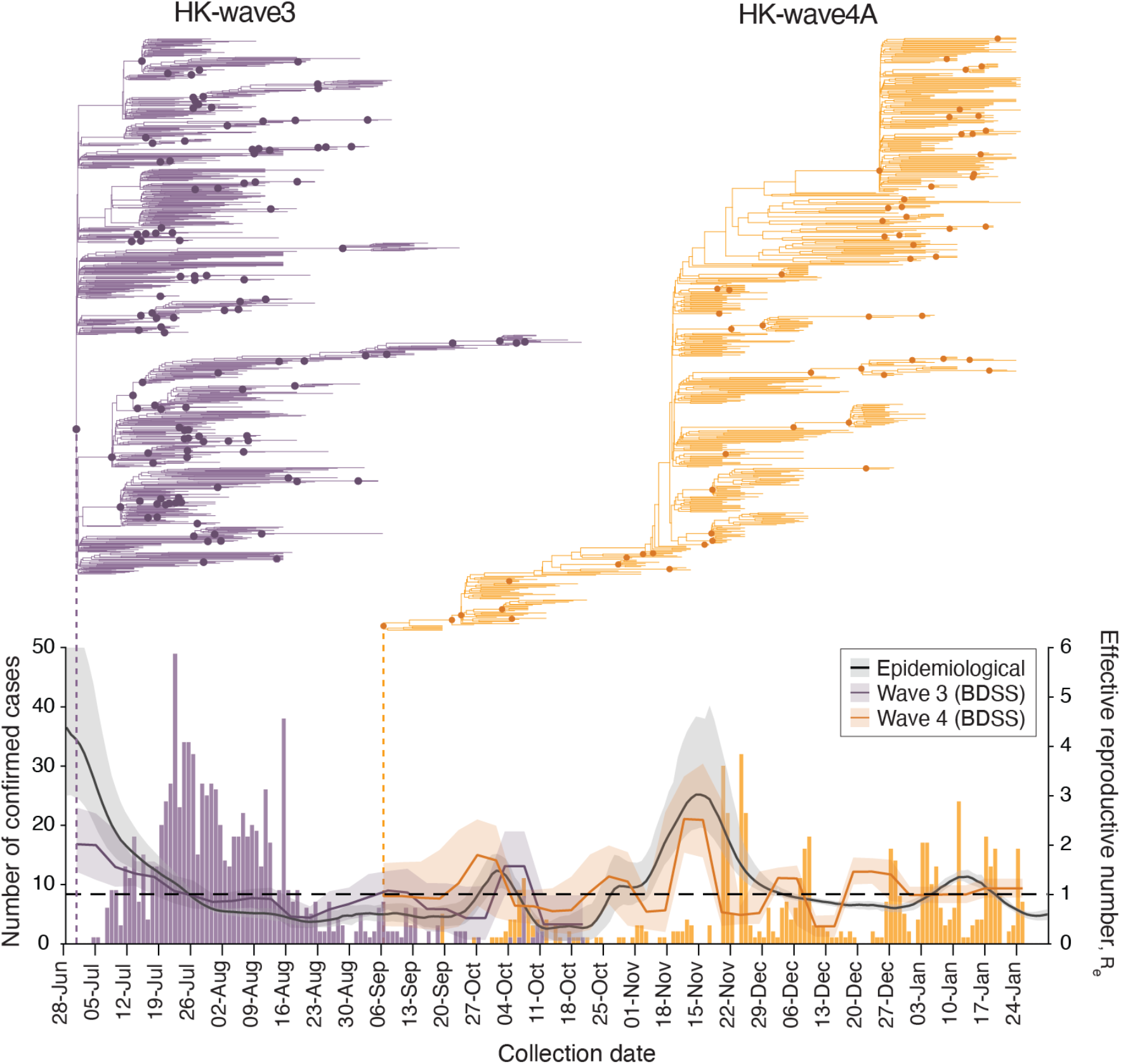
Phylodynamics of waves three and four in Hong Kong. Evolutionary relationships and effective reproduction number (R_e_(t)) of HK-wave3 (B.1.1.63) clade (purple) and HK-wave4A (B.1.36.27) clade (orange) estimated using tree-heights and sequenced incidence data. Purple and orange bars show the number of genomes by collection date. Black line shows the instantaneous effective reproduction number (R_t_), estimated based on infection dates inferred from the reported dates of symptom onset (or dates of confirmation for asymptomatic cases).

HK-wave4A (mean tMRCA = 6-September-2020, 95% HPD 5–7 September-2020) continued to circulate for several months despite level 5 stringency (**Fig. 3**). In contrast to HK-wave3, a rapid expansion did not occur upon emergence. Instead, the HK-wave4A dynamics resembled the pattern of a propagated epidemic curve, with R_e_ increasing with each subsequent peak during September and November, achieving a high of R_e_ ∼3 during mid to late November; R_e_ fluctuated around 1 following this period. The structure of the phylogenetic tree suggests continual elimination of viral lineages with intermittent expansion coinciding with a superspreading event involving 28 dancing and singing venues in different districts of Hong Kong, resulting in a group of 732 cases epidemiologically linked to this superspreading event. Epidemiological data shows that up to 23.6% of genomes during this wave were related to social clusters (**table S5**) which is significantly higher than the HK-wave3 counterpart (p < 0.001, chi-square test). Human mobility levels inferred from Octopus, a smart card payment system used by >98% of the Hong Kong population aged 16 to 65 (*15*), showed adult and elderly mobility within Hong Kong resumed to pre-pandemic levels during the early stage of wave four in early to mid-November 2020 (∼100% of the average level during 1–15 January 2020) (**fig. S5**). Taken together, our results suggest that community measures reduce R_e_, however prolonged community measures decrease the probability of lineage termination due to fatigue arising in the population (*16, 17*).

The instantaneous effective reproductive numbers (R_t_) of local cases are consistent with R_e_ of the two major transmission lineages during waves three and four (**Fig. 3**). During the short period of lineage co-circulation (**Fig. 3**), the relative reproductive number of HK-wave3 estimated using a relative fitness inference framework (*15*) was higher (mean = 1.28, credible interval [CrI] 0.88-1.93) when compared with HK-wave4A, though the difference in transmissibility was not significant (CI across 1.0). However, these estimates of comparative transmissibility may be affected by stochasticity, such as the occurrence of superspreading events as revealed through sequencing.

## Within-host genetic variation determined by genetic background

By analyzing deep-sequence data from confirmed donor and recipient pairs using a beta-binomial statistical framework (*18*) we estimated the number of virions required to initiate infection was between one to three (**Fig. 4A, Materials and Methods** and **table S8**). This is consistent with data from transmission pairs estimated in UK and Austria (one to eight in UK and one to three in Austria (*19, 20*)) as well as between cats (two to five virions (*21*)), showing SARS-CoV-2 transmission bottleneck may be universally small. To understand the effects of strong transmission bottlenecks on within-host diversity, we estimated Jaccard distances between sample pairs and found that minor variants were not significantly different between established transmission pairs and epidemiologically clustered samples (**Fig. 4B**) but were significantly different between samples without epidemiological links. These results show that the SARS-CoV-2 within-host genetic variation is non-random and determined by genomic differences (i.e. consensus sequence), shedding new light on SARS-CoV-2 evolution.

**Fig. 4.**
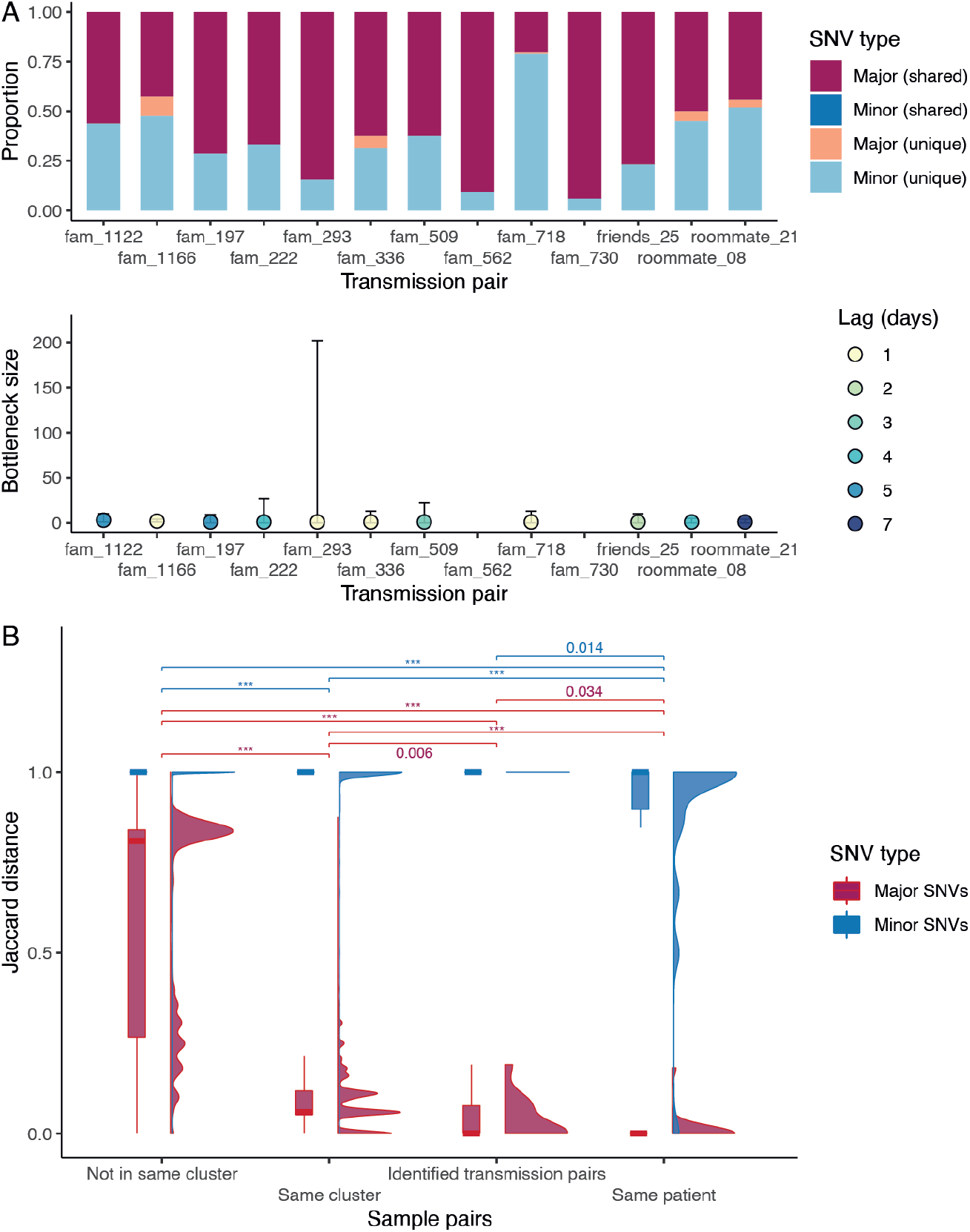
Transmission bottleneck and mutation profiles between cases. **(A)** Proportion of shared mutations and transmission bottleneck size estimations (with 95% confidence intervals) between samples from established transmission pairs. The estimates for transmission pairs fam_562 and fam_730 are not available due to a limited number of SNVs in the recipients’ samples. **(B)** Jaccard distance of the major and minor SNVs between different types of sample pairs. The distribution is shown in both boxplot and density plot. The between-group differences were tested by Wilcoxon tests, and p values < 0.05 are shown (p values < 0.001 are labelled as ***).

## Conclusions

Sequencing allowed us to investigate the introduction and circulation pattens of SARS-CoV-2 transmission lineages under the elimination strategy in Hong Kong. Border control measures averted numerous introductions, and community outbreaks were typically associated with cryptic virus circulation during less stringent periods and expansion through superspreading events. Heightened control measures eliminated most transmission chains in the local community, but the proportion of social transmission during wave four was significantly higher than that of wave three, indicating control was less effective when prolonged. Although public health and social measures were promptly lifted when community cases could be traced and controlled, new waves continued as a result of new introductions rather than resurgence of previously circulating viruses. This shows that contact tracing was efficient, but averting outbreaks from new introductions requires heightened border control and enhanced community surveillance during periods of lower control level stringency.

## Data Availability

Hong Kong SARS-CoV-2 genome sequences and associated metadata is deposited at gisaid.org. All anonymized data, sequence accession numbers, code, and analysis files are available in supplementary materials and in GitHub repository (https://github.com/hku-sph-covid-19-genomics-consortium/hk-sars-cov-2-genomic-epidemiology).

https://github.com/hku-sph-covid-19-genomics-consortium/hk-sars-cov-2-genomic-epidemiology

## Acknowledgments

We gratefully acknowledge the staff from the originating laboratories responsible for obtaining the specimens and from the submitting laboratories where the genome data were generated and shared via GISAID. We thank Octopus Cards Limited for providing aggregate data of passenger numbers by public transportation means and aggregate data of transactions by retail categories for the research. We acknowledge the technical support provided by colleagues from the Centre for PanorOmic Sciences of the University of Hong Kong. We also acknowledge the Centre for Health Protection of the Department of Health for providing epidemiological data for the study. The computations were performed using research computing facilities offered by Information Technology Services, the University of Hong Kong. The funding bodies had no role in the design of the study, the collection, analysis, and interpretation of data, or writing of the manuscript.

## Author contributions

Study Design: LLMP, DV

Data curation: HG, RX, DCA, DNCT, KME

Genome sequencing group: LLMP, JSMP, DKC, HG, LDJC, SSYC, SG, PK, DYMN, GYZL, CKCW

Phylodynamic analysis group: DV, TTYL, DCA, HG, RX, KSML, JL-HT Visualization: HG, RX, DCA, TTYL, DV

Supervision: LLMP, DV, BJC, JSMP, TTYL, JTKW

Writing – original draft preparation: DV, RX, DCA, HG, TTYL

Writing – review & editing: LLMP, DV, DCA, TTYL, KME, JSMP, BJC, GML

## Supplementary Materials

### Materials and Methods

#### SARS-CoV-2 data from Hong Kong

De-identified saliva or nasopharyngeal samples positive for SARS-CoV-2 by real time-polymerase chain reaction (RT-PCR), along with epidemiological information including onset date, report date and contact history for individual cases were obtained from the Centre for Health Protection, Hong Kong.

#### Genomic sequencing of SARS-CoV-2

A total of 1,753 laboratory-confirmed samples were collected from 1,733 RT-PCR confirmed cases from 22-June-2020 to 26-January-2021. Virus genome was reverse transcribed with primers targeting different regions of the viral genome, published in (*22*). The synthesized cDNA was then subjected to multiple overlapping 2kb PCRs for full-genome amplification. PCR amplicons obtained from the same specimen were pooled and sequenced using Nova sequencing platform (PE150, Illumina). Sequencing library was prepared by Nextera XT. The base calling of raw read signal and demultiplexing of reads by different samples were performed using Bcl2Fastq (Illumina). A reference-based re-sequencing strategy was applied in analyzing the NGS data. Specifically, the raw FASTQ reads were assembled and mapped to the SARS-CoV-2 reference genome (Wuhan-Hu-1, GenBank: MN908947.3) using BWA mem2 (v.2.0pre2) (*23*). The consensus sequences for each sample were called as dominant bases at each position by samtools mpileup (v.1.11) (*24*) with minimum depth of 100 reads. Samples less than 27kb in length (excluding gaps) were excluded from downstream analysis. The head and tail 100nt bases of all generated consensus sequences were masked. We also masked another 10 sites located in PCR primer binding regions and observed to be variant (≥ 3% allele frequency) in 1% or more HK samples (**table S9**). The same masking strategy was also applied in phylodynamics analysis, variant calling, and bottleneck estimation. The average sequencing depth (number of mapped reads) at each nucleotide position that was retained ranged from ∼10,000 to ∼100,000 (**fig. S6**).

There were 16 patients from which samples were collected or sequenced at multiple time points. Twelve samples from 6 patients were sequenced in duplicate, and 21 samples from 10 patients were collected sequentially. One representative sample for each of the 16 patients was selected based on genome coverage and average sequencing depth. In total, 1,601 representative samples met quality control standards. All 1,601 consensus sequences from Hong Kong, as well as 298 additional consensus sequences from the first two waves were included in the phylogenetic analysis. Sequences from regions outside Hong Kong were retrieved from the GISAID database (total 399,124 sequences, accessed 16-February-2021, detailed accession numbers and acknowledgement information in **data S4**).

#### Phylogenetic analysis of SARS-CoV-2 in Hong Kong

Hong Kong sequences were analysed with a global SARS-CoV-2 genome alignment obtained from GISAID (accessed 16-February-2021). For each Hong Kong sequence, the three most similar global sequences (evaluated by p distance excluding gaps, n = 385), as well as the earliest sampled sequence (n = 1,279) from each PANGO lineage (accessed 07-May-2021) (*11*) were selected. After removing repetitive sequences and trimming masked sites, data quality was evaluated using a root-to-tip regression analysis in TempEst (v.1.5.3) (*25*), resulting in a final set of 3,437 sequences. Maximum likelihood (ML) phylogenies were estimated using IQ-TREE (v.2) (*26*), employing the best-fit nucleotide substitution model with Wuhan-Hu-1 (GenBank: MN908947.3) as the outgroup and dated by least square dating (LSD2) (*27*). Branch support was estimated using ultrafast bootstrap approximation (UFBoot) and SH-like approximate likelihood ratio test (SH-aLRT), and for nodes of interest with <50% support, we examined their stability through multiple iterative runs using the best-fit nucleotide substitution model. Internal branches with zero-length were preserved for dating by setting parameter *l* as -1. SARS-CoV-2 sequences from Hong Kong were classified based on the dynamic PANGO nomenclature system (https://github.com/cov-lineages/pangolin, v.2.3.9, 23-April-2021) (*11*) and confirmed using a ML analysis.

#### Phylodynamics of Hong Kong waves

To identify monophyletic clusters of SARS-CoV-2 lineages in Hong Kong, Bayesian molecular clock phylogenetic analysis pipeline proposed by Plessis *et al*. (*28*) were implemented. In this study, the data tree and starting tree were applied to a ML tree generated by IQ-TREE (v.2) (*26*) with LSD2 (*27*). Time-scaled phylogenies were generated using the strict clock model with 0.001 substitutions per site per year which is within 95% credible interval of SARS-CoV-2 temporal signal (*29*), the Skygrid model (*30*) with 61 grid points and a Laplace root-height prior with mean equal to the dated-ML tree estimated by IQ-TREE (v.2) (*26*) and scale is set to 20% of mean. To improve computational efficiency, two largest local monophyletic clades in wave three (HK-wave3, n = 902) and wave four (HK-wave4A, n = 552) from ML tree were subsampled to 100 and 65 sequences by 5 earliest cases, 5 latest cases and 10% of the remaining randomly selected, respectively. We ran nine MCMC chains of 100 million, sampling every 1,000 steps and discarding 10% as burn-in. As there are no collapsed internal branches in this study, only uncertainty in branch durations were estimated by MCMC. From the approach described in Geoghegan *et al*. (*31*), we used the R package “NELSI” (*32*) to identify classify all monophyletic lineages, including singletons, and to estimate the delay in lineage detection following importation as well as the duration of circulation, given a set of 8,000 posterior trees. It’s notable that there are two global sequences from Japan (EPI_ISL_591420 and EPI_ISL_721612) present in the HK-wave3 clade. However, these two Japan cases had travel history to the Philippines (similar to the early HK-wave3 imported cases), and were quarantined when landed in Japan, suggesting that they are unlikely to have caused an introduction in Hong Kong. These two cases were therefore excluded when defining the HK-wave3 clade.

For all samples of HK-wave3 and HK-wave4A, we used the birth-death skyline serial (BDSS) model (*14*) implemented in BEAST (v.2.6.3) (*33*) to infer the time of origin (tOrigin), time of most recent common ancestor (tMRCA) and temporal variations (piecewise fashion over 12-15 equidistant intervals) in the effective reproductive number denoted as R_t_. A non-informative prior for tOrigin was used with a the lower bound set to 1-January-2020. The HKY + G4 nucleotide substitution model and an uncorrelated relaxed molecular clock model with lognormal rate distribution (UCLN) (*34*) were used. The sampling proportion was given a uniform distribution as prior with the upper bound at the empirical ratio of the number of sequences to the number of reported cases. MCMC chains were run for 600 million and 800 million steps and sampled every 2,000 and 10,000 steps for the lineages HK-wave3 (B.1.1.63) and HK-wave4A (B.1.36.27) respectively, with the initial 10% discarded as burn-in. This resulted in a final total of 270,000 and 72,000 sampled states. Mixing of the MCMC chain was inspected using Tracer (v1.7.1) (*35*) to ensure an effective sample size (ESS) of >200 for each parameter. Change in the effective reproductive number over time after the estimated tMRCA was plotted using R package “bdskytools” (https://github.com/laduplessis/bdskytools). Since by definition there are no sequences between tMRCA and the estimated tOrigin, the effective reproductive number (R_e_) was assumed to remain constant in this period. This assumption was incorporated in the default birth-death model using the package TreeSlicer in BEAST2.

#### Human mobility in Hong Kong using Octopus data

We used digital transactions made on Octopus cards, ubiquitously used by the Hong Kong population for daily public transport and small retail payments (https://www.octopus.com.hk/tc/consumer/index.html), to obtain changes in mobility during 2020–2021 among cards classified as children, students, adults and elderly (**fig. S5**).

#### Analysis of within-host genetic variation and transmission bottleneck

Deep sequencing SARS-CoV-2 samples in the United Kingdom and Austria has shown that the within-host genetic diversity (iSNV) is low with a narrow bottleneck during transmission (*19, 20*). Within-host genetic variation (iSNV) depends on intra-host virus evolution and transmission bottleneck size. To determine the baseline similarity of within-host genetic diversity, we prepared two sets of samples as controls: (a) six samples were sequenced in duplicate to account for uncertainty arising from sequencing; and (b) twenty-one samples collected from ten individuals. To examine the dynamics of mutations related to transmission events, we identified 13 transmission pairs (donor and recipient) that were directly linked with symptom onset varying by 1 to 7 days.

Single nucleotide variants (SNVs) in deep-sequence data were identified using three different variant callers, freebayes (v.1.3.2) (*36*), VarDict (v.1.82) (*37*) and LoFreq (v.2.15) (*38*). To attain a robust variant calling result, only SNVs detected by at least two different variant callers were analyzed in this study (*8*). SNVs with a minimum depth of 100 reads, minimum frequency of 3%, and detected by at least two different variant callers were retained for further analysis. Parameters and scripts for this pipeline are described in https://github.com/hku-sph-covid-19-genomics-consortium/hk-sars-cov-2-genomic-epidemiology. Gene annotations of the SNVs were based on **table S10**. To understand the uncertainty in iSNV due to the sequencing protocol, we sequenced six samples in duplicate. While detection of major variants (consensus) was consistent, detection of iSNVs may vary between sequencing runs. For example, while iSNVs of case 10 were shared between sequencing runs, iSNVs of case 15 were mutually exclusive (**fig. S7**). Similarly, when comparing iSNV of 21 samples collected from 10 individuals, major variants in samples from the same patient remained identical but their iSNVs varied. A similar pattern was observed in transmission pairs, where most of the major variants are shared but some of the minor variants are unique (**Fig. 4A**), which can be explained by a narrow transmission bottleneck.

The statistical framework for estimating the transmission bottleneck size between identified transmission pairs was introduced in (*18*). It was based on a beta-binomial method which models the number of transmitted virions from donor to the recipient. Because of the high sequencing depth of the data, the minimum variant calling threshold was set to 0.03 with a minimum depth of 100 reads. Bottleneck size estimates were calculated by maximum likelihood analysis comparing the allele frequency of variants passing threshold between samples. The 95% confidence intervals were calculated using a likelihood ratio test. To identify similarity of SNVs between samples we used the Jaccard distance, defined as one minus the proportion of intersection between two samples divided by the proportion of their union.

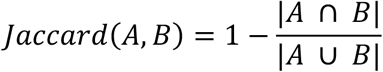

SARS-Cov-2 sequences from Hong Kong contained within-patient variation in 12,859 sites of the genome when compared to the SARS-CoV-2 reference strain Wuhan-Hu-1 (GenBank: MN908947.3). 37.2% of the sites (n=4,779) contained mutations in more than one sample. High frequency of variation (in >100 Hong Kong sequences) was observed in 30 sites (**table S6**). The spectrum of allele frequencies (**fig. S8**) showed that over 90% of the variants had allele frequency ≥95% or ≤10%. Mutation hotspots were detected in four sites of the genome with a greater frequency of intra-host single nucleotide variation (iSNVs). These mutations were sporadically distributed across the phylogeny and present in ∼20 sequences from the GISAID data, which may suggest potential homoplasy. We observed two variants unique to Hong Kong, C5812T and G25785T, which were detected in separate phylogenetic clusters of local Hong Kong cases. The C5812T mutation was identified in two sperate clusters in the fourth wave. While the C5812T mutation in one cluster likely descended from a local ancestral case, the mutation in the earlier cluster may have been imported. Similarly, G25785T was found in both the third and fourth waves, and mutations in at least one cluster likely originated from local cases. Some of the low frequency SNVs (frequency <5%, shown in the low peaks to the bottom of **fig. S8** and **table S7**) commonly occurred in global context. For example, the G28883C (G205R in nucleocapsid) and C22227T (A665V in spike) mutations were found in 38.09% and 21.49% of the global cases, however they were only seen in 1.56% and 0.7% of the Hong Kong cases respectively.

#### Estimation of the instantaneous effective reproductive number (*R*_*t*_)

The instantaneous effective reproduction number *R*_*t*_ is defined as the average number of secondary cases generated by cases on day *t*. If *R*_*t*_ > 1 the epidemic is expanding at time *t*, whereas *R*_*t*_ < 1 indicates that the epidemic size is shrinking at time *t*. The transmissibility of imported and local cases was expected to be very different because intensive non-pharmaceutical interventions had been imposed on travelers arriving from COVID-19 affected regions since January 2020. Hence, we only included cases from the following three categories into the computation of *R*_*t*_, i.e., local case and epidemiologically linked with local cases defined by the Centre for Health Protection (CHP, https://www.coronavirus.gov.hk/eng/index.html).

Since the epidemic curves provided by CHP were based on the dates of symptom onset or dates of confirmation, we used a deconvolution-based method to reconstruct the COVID-19 epidemic curves by dates of infection (*39, 40*). We assumed that the incubation period was Gamma with mean and standard deviation of 6.5 and 2.6 days (*41*), and that the distribution of the time between symptom onset and case confirmation was Gamma with mean and standard deviation of 4.3 and 3.2 days. For asymptomatic cases, we assumed they shared the same distribution of the time between infection and case confirmation with the symptomatic cases. We then computed *R*_*t*_ for local cases only from the respective epidemic curves using the “*EpiEstim*” (*42*) R package (**Fig. 3**).

#### Estimation of the relative reproductive number of HK-wave3 compared with HK-wave4A

We defined the comparative transmissibility of any two lineages as the relative reproductive number, i.e., the ratio of their basic reproductive numbers. We extended a previous competition transmission model (*43, 44*) of two viruses and applied the fitness inference framework to the sequence data collected in Hong Kong during the cocirculation period of HK-wave3 and HK-wave4A clades (between 19-September and 21-October-2020, **Fig. 3**). We assumed the two clades shared the same generation time distribution which can be approximated by the serial interval distribution estimated in Leung *et al*. (*45*) (i.e., Gamma distribution with mean and standard deviation of 5.2 and 1.7 days). The inference framework incorporates both incidence and genotype frequency data that reflect the local comparative transmissibility of cocirculating lineages.

---

**Fig. S1.**
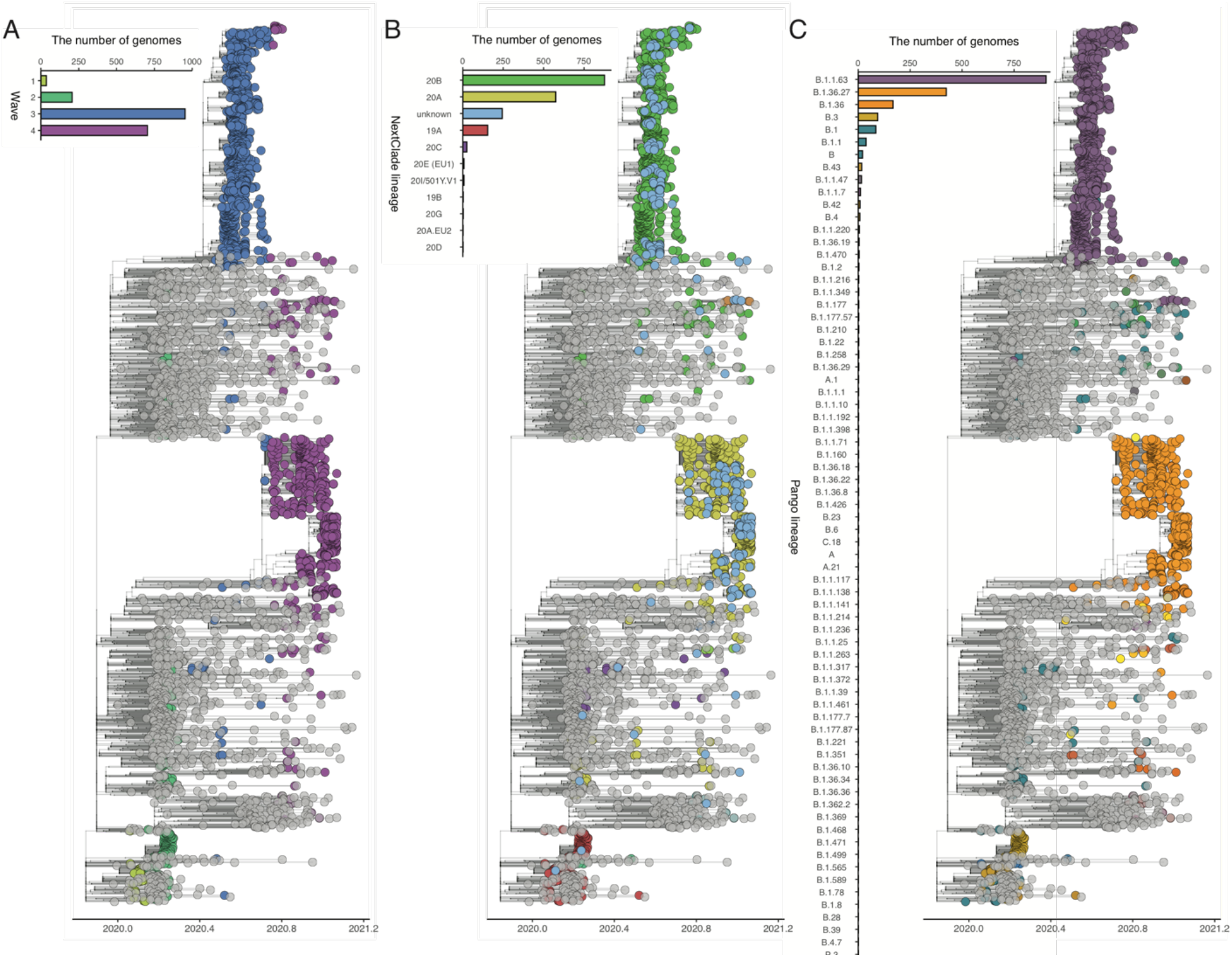
Time-scaled maximum-likelihood phylogeny of SARS-CoV2 using IQ-TREE (v.2) (*26*). The tree colored by (**A**) pandemic waves in Hong Kong, (**B**) Nextclade, and (**C**) PANGO classification, respectively. Global sequences are shown in grey.

**Fig. S2.**
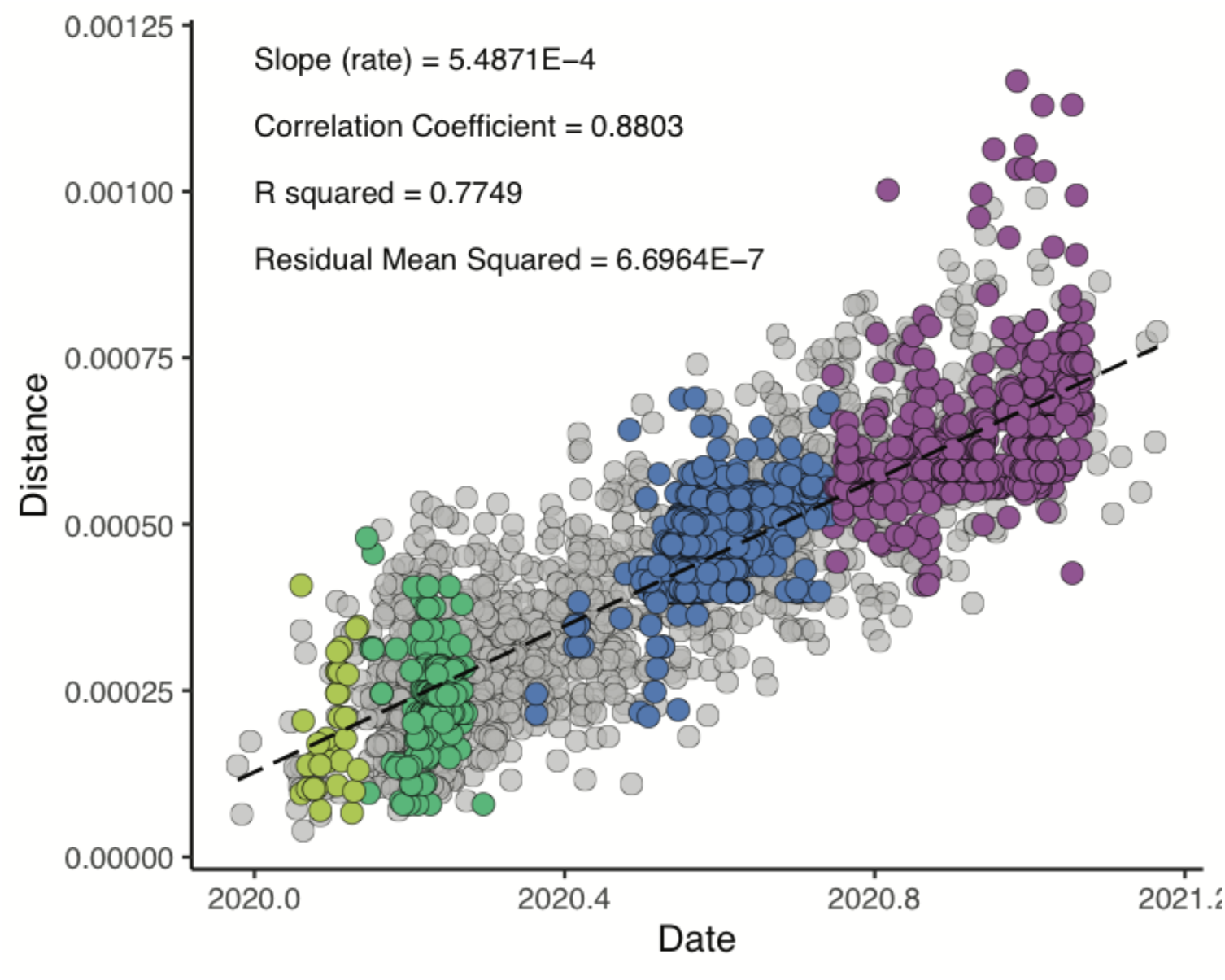
Root-to-tip regression analysis was performed in TempEst v.1.5.3 (*25*). Colours indicate pandemic waves in Hong Kong as in **fig. S1 (A)**.

**Fig. S3.**
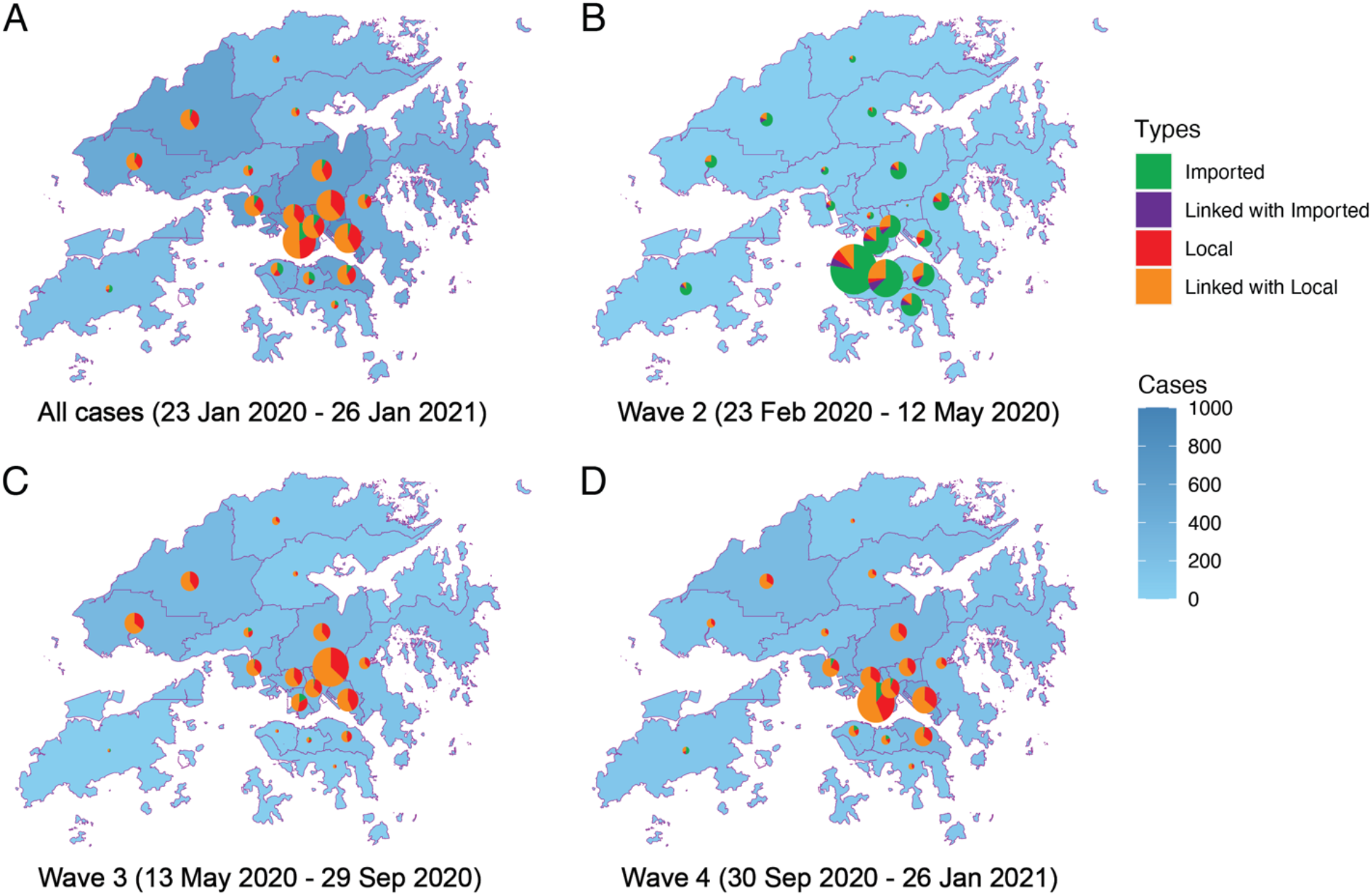
Geography of SARS-CoV-2 cases in Hong Kong. (**A**) Map of Hong Kong’s 18 districts shaded by the number of laboratory-confirmed cases of SARS-CoV-2. Pie charts divided by transmission types. Pie chart size reflects the number of laboratory-confirmed cases in this district (ratio of cases and radius: 1/50,000). (**B**) same as (**A**) based on second wave (ratio of cases and radius in pie charts: 1/5,000). (**C**) same as (**A**) based on third wave (ratio of cases and radius in pie charts: 1/25,000). (**D**) same as (**A**) based on fourth wave (ratio of cases and radius in pie charts: 1/30,000).

**Fig. S4.**
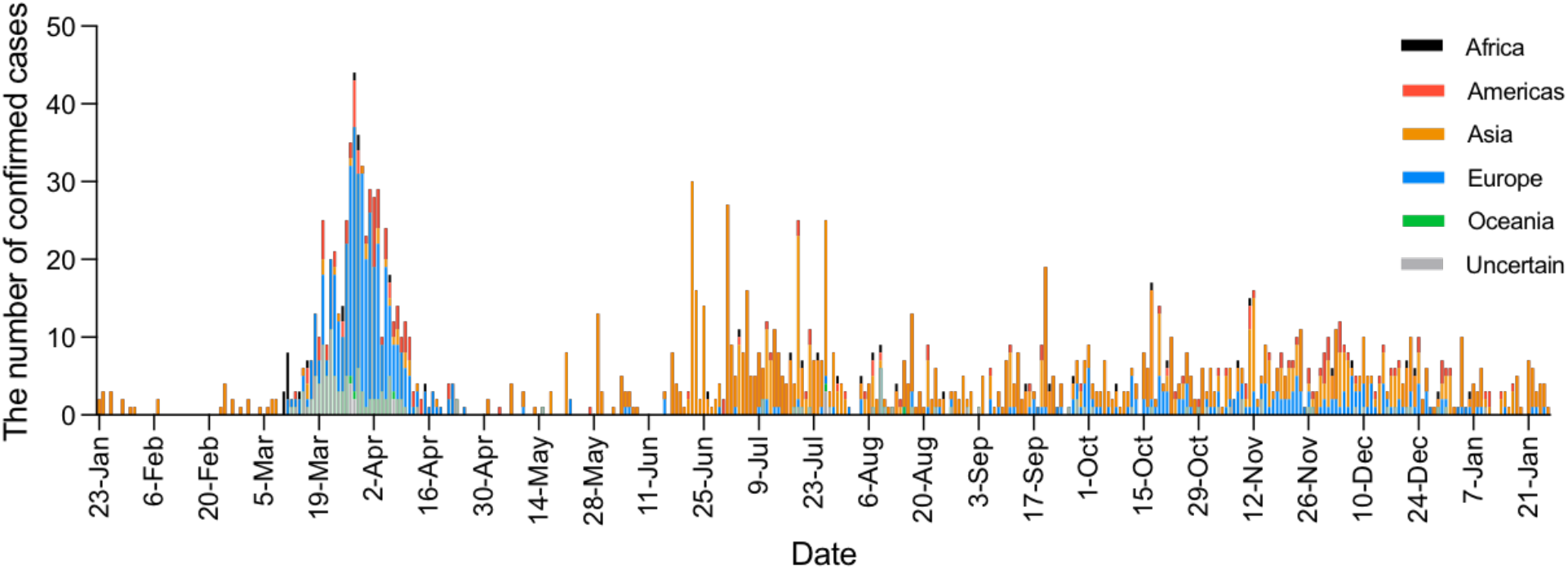
SARS-CoV-2 imported cases colored by continent of origin.

**Fig. S5.**
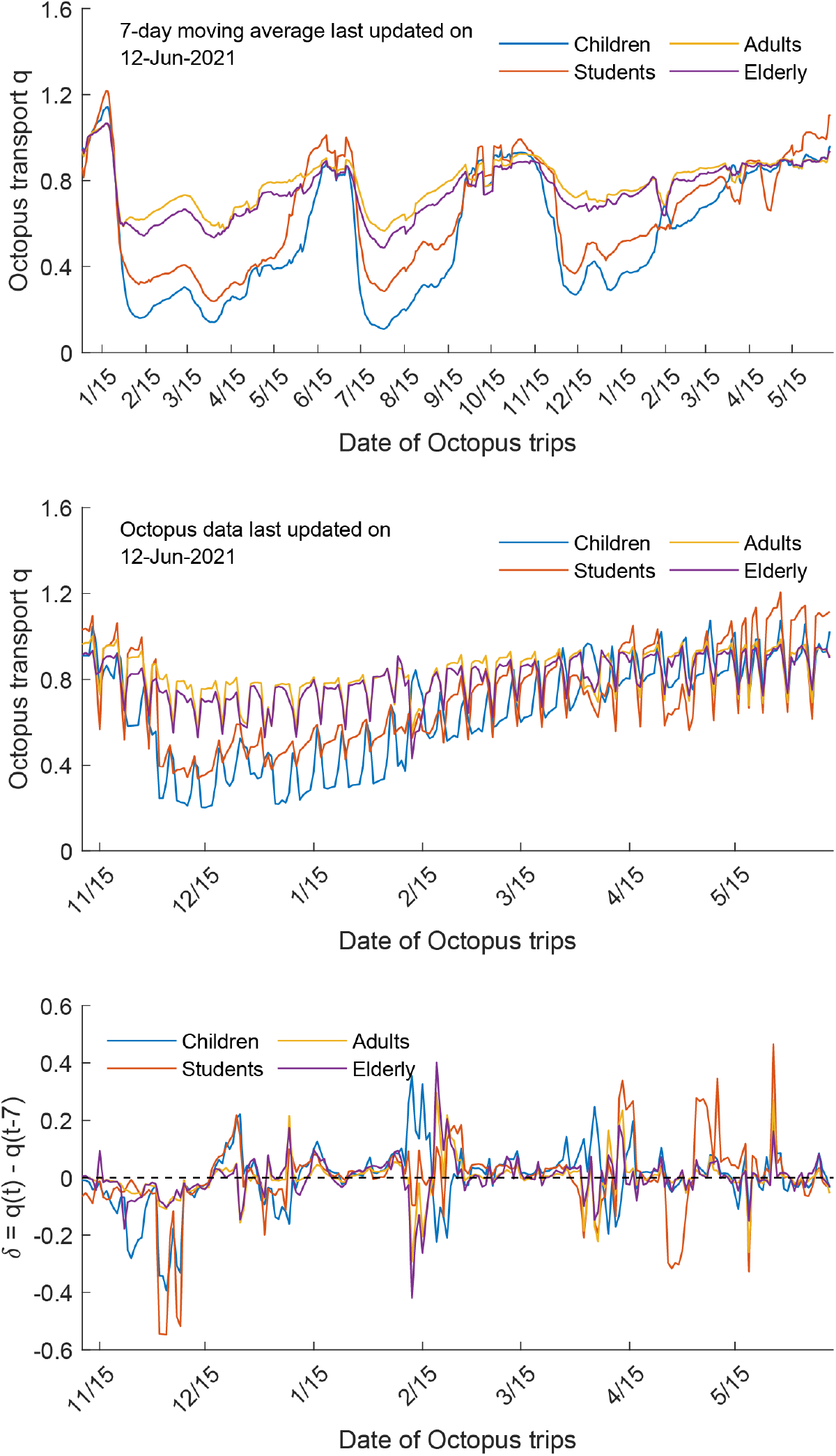
Octopus mobility data during pre-pandemic period and early period of wave four in Hong Kong.

**Fig. S6.**
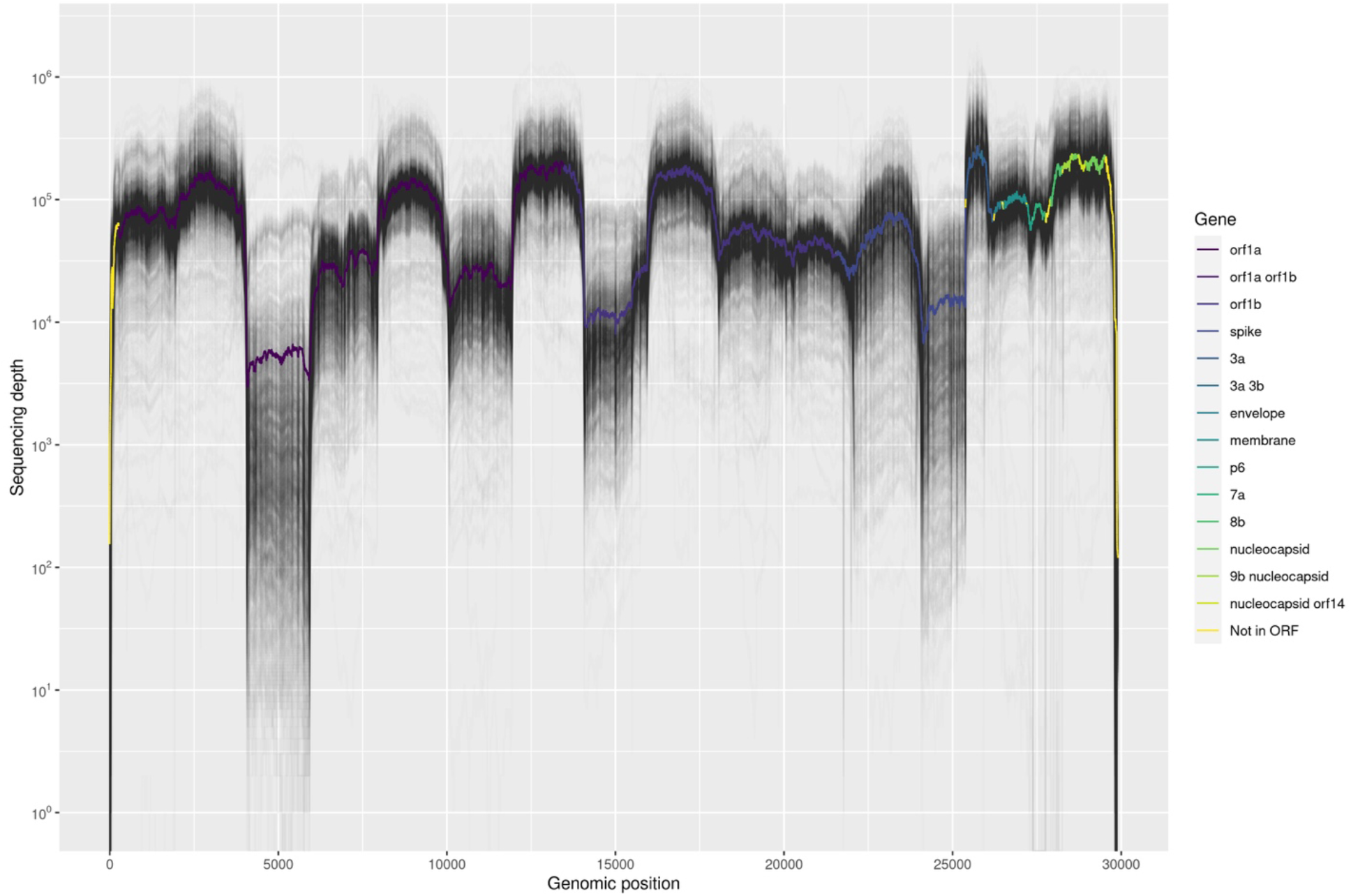
Sequencing depth of NGS data across the genome. The colored line represents the average read depth at each genomic position.

**Fig. S7.**
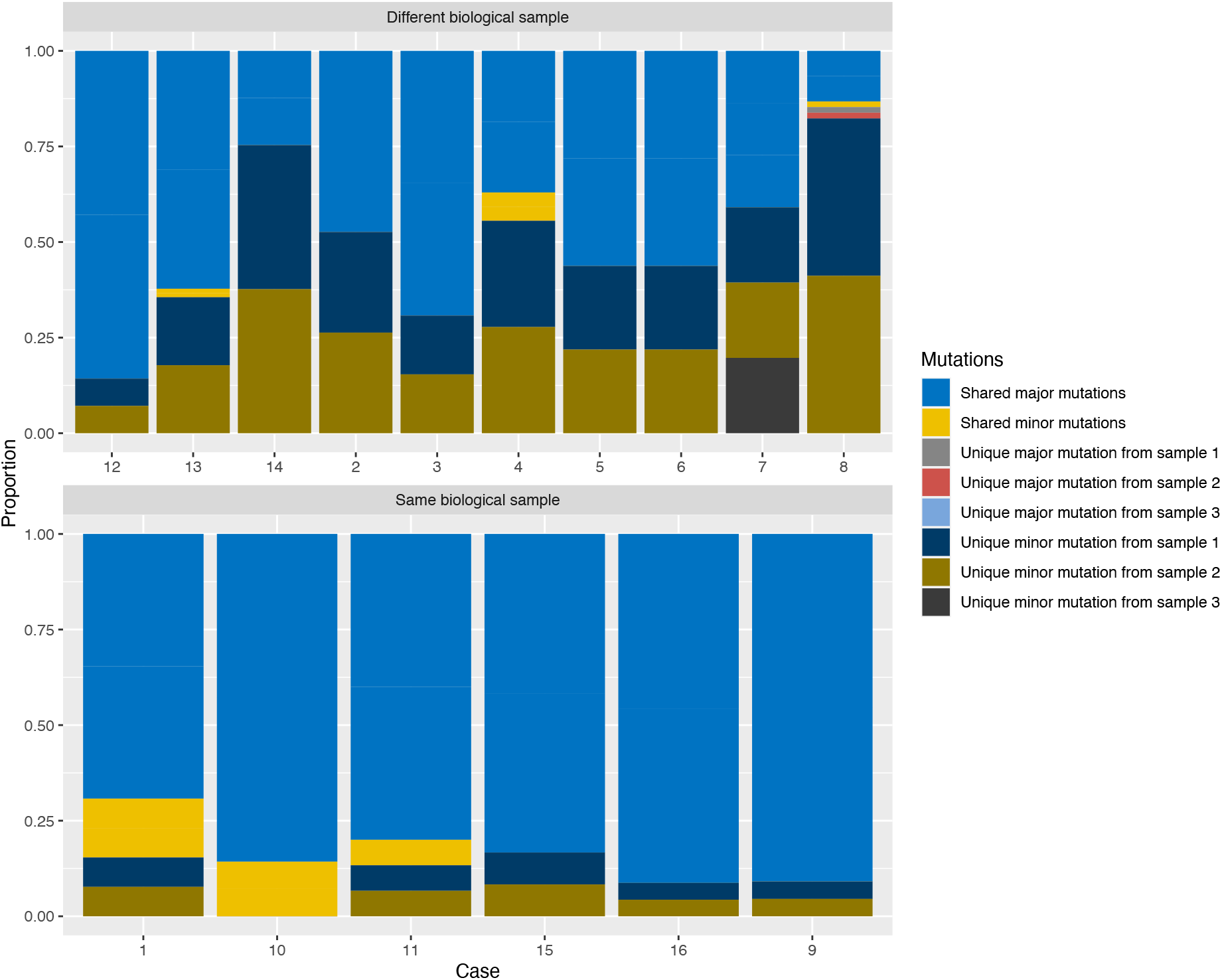
Proportion of shared mutations between samples from same individuals.

**Fig. S8.**
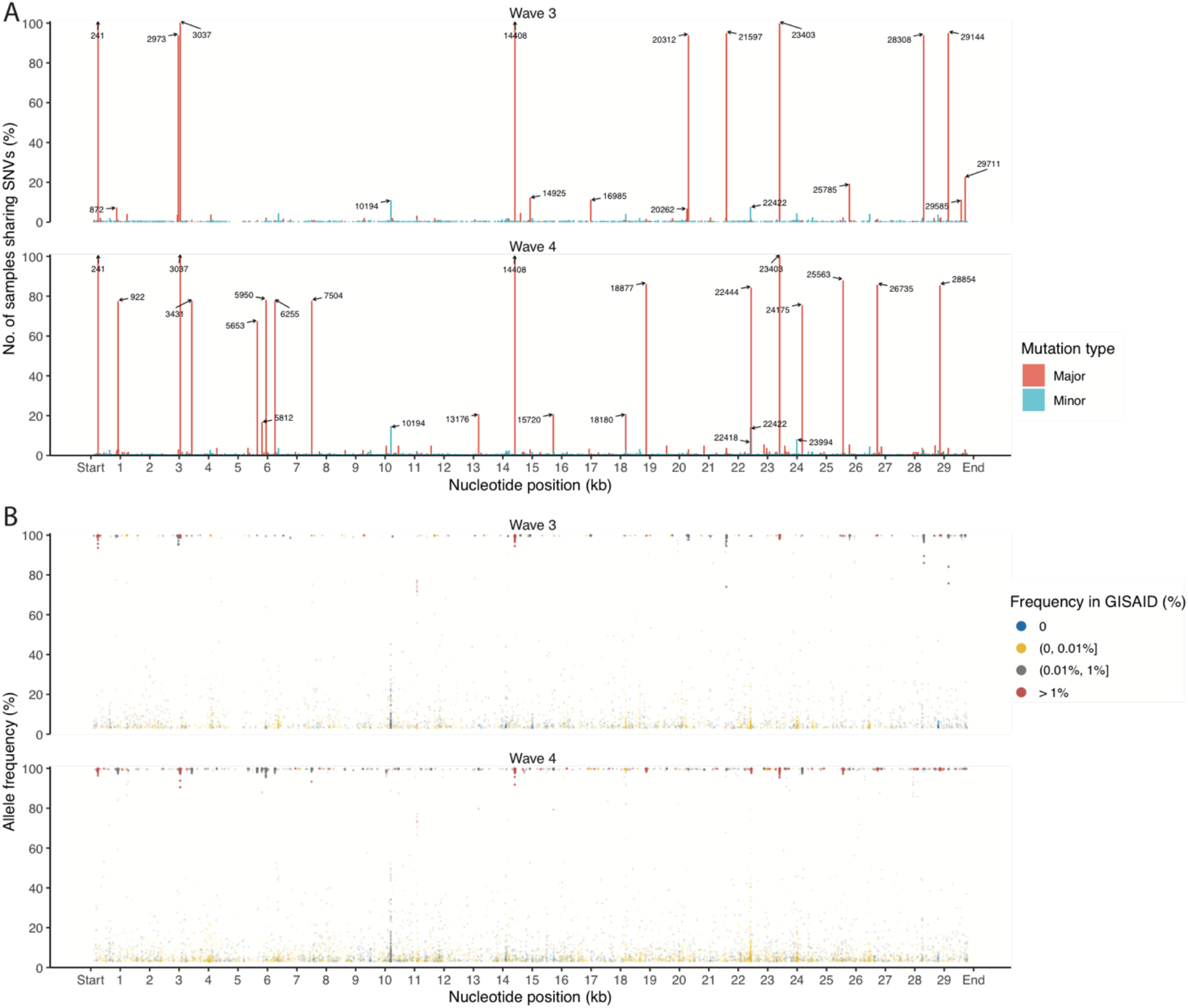
Frequencies and dynamics of SNVs in Hong Kong. (**A**) Percentage of samples that share mutations in third and fourth wave epidemics. The color codes for major and minor mutations at different genomic sites. High-frequency variant sites (n = 36, identified in at least 5% of the respective samples) are labeled. (**B**) Relative mutation frequency (allele frequency) of SNVs. The SNVs are colored by the frequencies of mutations reported in GISIAD dataset.

**Table S1.** Cases and genome sequences from waves of SARS-CoV-2 in Hong Kong.

| Wave | Duration | Peak of wave (no. of cases on peak day) | Travel-related cases <sup>b</sup> | Community cases <sup>c</sup> | Total confirmed cases | Genomes sequenced | Changes in stringency <sup>d</sup> |
| --- | --- | --- | --- | --- | --- | --- | --- |
| 1 | 23-Jan to 22-Feb | 9-Feb (10 cases) | 17 | 53 | 70 | 36 | 1-2-3 |
| 2 | 23-Feb to 12-May | 27-Mar (65 cases) | 705 | 273 | 978 | 206 | 3-2-3-4 |
| 3 | 13-May to 29-Sep | 30-Jul (149 cases) | 647 | 3385 | 4032 | 953 | 4-3-2-4-3 |
| 4<sup>a</sup> | 30-Sep to 26-Jan 2021 | 29-Nov (115 cases) | 629 | 4515 | 5144 | 704 | 3-4-5 |

<sup>a</sup>wave four is summarized until 26 Jan 2021 for this study. <sup>b</sup> imported cases only. <sup>c</sup> epidemiologically linked with imported cases, local cases, and epidemiologically linked with local cases. <sup>d</sup>A-B denotes the stringency index level (based on Oxford COVID-19 Government Response Tracker, See Methods) change from A to B (e.g., 1-2-3 means that the stringency index level was increased from 1 to 2 to 3).

**Table S2.** Genome sequencing and population statistics of Hong Kong districts.

| Districts | SARS-COV-2 sample size <sup>a</sup> | Sequenced <sup>b</sup> | Latitude <sup>c</sup> | Longitude <sup>c</sup> | Population size <sup>d</sup> | Population density <sup>d</sup> (people per km<sup>2</sup>) | Median monthly household income <sup>d</sup> (HK\$) |
| --- | --- | --- | --- | --- | --- | --- | --- |
| Central and Western | 365 | 58 | 22.282150 | 114.156880 | 240,500 | 19,171 | 41,400 |
| Eastern | 548 | 81 | 22.284030 | 114.224220 | 545,600 | 30,336 | 34,300 |
| Southern | 219 | 42 | 22.246760 | 114.174134 | 264,600 | 6,812 | 32,800 |
| Wan Chai | 348 | 76 | 22.279680 | 114.171680 | 178,400 | 16,973 | 44,100 |
| Sham Shui Po | 626 | 126 | 22.330700 | 114.162163 | 416,500 | 44,517 | 24,300 |
| Kowloon City | 646 | 140 | 22.328290 | 114.191490 | 419,900 | 41,919 | 30,000 |
| Kwun Tong | 804 | 153 | 22.313260 | 114.225810 | 688,500 | 61,075 | 22,500 |
| Wong Tai Sin | 841 | 132 | 22.342140 | 114.195830 | 416,100 | 44,729 | 25,500 |
| Yau Tsim Mong | 977 | 216 | 22.321320 | 114.172580 | 329,900 | 47,177 | 30,000 |
| Islands | 203 | 38 | 22.210370 | 114.028800 | 186,500 | 1,054 | 28,400 |
| Kwai Tsing | 566 | 124 | 22.354880 | 114.084010 | 502,400 | 21,528 | 24,700 |
| North | 207 | 43 | 22.494711 | 114.138123 | 314,100 | 2,301 | 30,400 |
| Sai Kung | 400 | 78 | 22.381540 | 114.270393 | 472,500 | 3,645 | 36,500 |
| Sha Tin | 601 | 99 | 22.387159 | 114.195229 | 688,100 | 10,014 | 29,700 |
| Tai Po | 246 | 50 | 22.450840 | 114.164223 | 306,800 | 2,254 | 25,800 |
| Tsuen Wan | 288 | 66 | 22.374630 | 114.115097 | 311,800 | 5,034 | 32,600 |
| Tuen Mun | 473 | 127 | 22.396910 | 113.974411 | 495,100 | 5,964 | 25,000 |
| Yuen Long | 553 | 134 | 22.445570 | 114.022293 | 645,000 | 4,711 | 27,000 |

<sup>a</sup> There are 1313 cases that have unknown location information. <sup>b</sup> There are 116 genomes that have unknown location information. <sup>c</sup> Data from https://www.latlong.net/. <sup>d</sup> Data from the Census and Statistics Department (Hong Kong) in 2019.

**Table S3.** Travel related cases and epidemiologically linked with imported cases within seven Hong Kong monophyletic clades (over 10 community cases).

| The number of samples within lineage | Travel related case ID | Report date | District | Origin <sup>a</sup> |
| --- | --- | --- | --- | --- |
| 902 | 349 | 2020-07-15 | Unknown | Philippines |
|  | 1156 | 2020-09-11 | Unknown | Philippines |
|  | 783 | 2020-08-02 | Eastern | Philippines |
| 552 | 1180 | 2020-09-20 | Kowloon City | Nepal |
|  | 1182 | 2020-09-20 | Yau Tsim Mong | Nepal |
|  | 1209 | 2020-10-05 | Yau Tsim Mong | Nepal |
|  | 1183 | 2020-09-20 | Yau Tsim Mong | Nepal |
|  | 1211 | 2020-10-06 | Yau Tsim Mong | Nepal |
|  | 1203 | 2020-10-04 | Yau Tsim Mong | Nepal |
|  | 1208 | 2020-10-05 | Yau Tsim Mong | Nepal |
|  | 1184 | 2020-09-20 | Yau Tsim Mong | Nepal |
|  | 1181 | 2020-09-20 | Kowloon City | Nepal |
| 92 | NA<sup>a</sup> |  |  |  |
| 33 | 1596 | 2020-12-24 | Central and Western | United Kingdom |
|  | 1591 | 2020-12-22 | Wan Chai | United Kingdom |
|  | 1592 | 2020-12-23 | Yau Tsim Mong | India |
|  | 1593 | 2020-12-23 | Yau Tsim Mong | India |
|  | 1590 | 2020-12-22 | Wan Chai | India |
|  | 1586 | 2020-12-21 | NA | India |
| 29 | 84 | 2020-03-21 | Yuen Long | United Kingdom |
|  | 91 | 2020-03-22 | Tsuen Wan | United Kingdom |
|  | 81 | 2020-03-20 | Eastern | United Kingdom |
|  | 68 | 2020-03-18 | Shatin | United Kingdom |
|  | 78 | 2020-03-20 | Eastern | United Kingdom |
|  | 111 | 2020-03-24 | Central and Western | United Kingdom |
|  | 93 | 2020-03-22 | Eastern | United Kingdom |
|  | 92 | 2020-03-22 | Eastern | United Kingdom |
|  | 77 | 2020-03-20 | Kwai Tsing | United Kingdom |
|  | 71 | 2020-03-20 | Central and Western | United Kingdom |
|  | 90 | 2020-03-22 | Tsuen Wan | United Kingdom |
|  | 82 | 2020-03-20 | Shatin | United Kingdom |
|  | 83 | 2020-03-21 | Yuen Long | United Kingdom |
| 19 | NA<sup>b</sup> |  |  |  |
| 16 | 256 | 2020-07-01 | Shatin | United States of America |

<sup>a</sup> This result is based on epidemiological data. <sup>b</sup> No travel-related cases.

**Table S4.** Distribution of PANGO lineages across HK-wave3 and HK-wave4A clades.

| HK clade | PANGO lineage | Number of genomes |
| --- | --- | --- |
| HK-wave3 | B.1.1.63 | 888 |
|  | B.1.1 | 5 |
|  | B.1.1.220 | 6 |
|  | B.1.1.192 | 2 |
|  | B.1.1.398 | 1 |
| HK-wave4A | B.1.36.27 | 424 |
|  | B.1.36 | 124 |
|  | B.1.36.29 | 2 |
|  | B.1.36.10 | 1 |
|  | B.1.36.34 | 1 |

**Table S5.** Sample numbers associated with different transmission settings in HK-wave3, HK-wave4A and all community cases in wave3 and wave4 based on epidemiological data.

| **Transmission setting** | **Wave3 community cases (epidemiological data)** | **Wave4 community cases (epidemiological data)** | **HK-wave3 clade (sequenced data)** | **HK-wave4A clade (sequenced data)** |
| --- | --- | --- | --- | --- |
| Social | 408 | 802 | 114 | 130 |
| Rche/Rchd <sup>a</sup> | 145 | 117 | 47 | 4 |
| Family/Roommate | 1800 | 2352 | 470 | 226 |
| Unknown/Sporadic | 661 | 763 | 149 | 84 |
| Work | 354 | 421 | 118 | 81 |
| Nosocomial | 17 | 60 | 4 | 27 |

<sup>a</sup> Residential care homes for the elderly and disabled.

**Table S6.** Major SNVs identified in third and fourth waves in Hong Kong.

| Position | Wave | Gene | Synonymous mutation | Mutation at nucleotide | Mutation at amino acid | Number of samples with major SNVs | Number of samples with iSNVs | Frequency in GISAID | Proportion in GISAID (%) |
| --- | --- | --- | --- | --- | --- | --- | --- | --- | --- |
| 241 | Wave 3 & 4 | Not in ORF | Not in ORF | C241T | NA | 1598 | 0 | 377531 | 94.59 |
| 3037 | Wave 3 & 4 | nsp3 | TRUE | C3037T | F106F | 1600 | 0 | 381081 | 95.48 |
| 10194 | Wave 3 & 4 | nsp5 | FALSE | A10194T\|G | E47V\|G | 2 | 392 | 43 | 0.01 |
| 14408 | Wave 3 & 4 | nsp12_2 | FALSE | C14408T | P314L | 1595 | 0 | 380926 | 95.44 |
| 22422 | Wave 3 & 4 | S | FALSE | A22422G\|T | D287G\|V | 5 | 322 | 28 | 0.01 |
| 23403 | Wave 3 & 4 | S | FALSE | A23403G | D614G | 1597 | 0 | 381443 | 95.57 |
| 872 | Wave 3 | nsp2 | FALSE | G872A\|T\|C | D23N\|Y\|H | 82 | 4 | 679 | 0.17 |
| 2973 | Wave 3 | nsp3 | FALSE | C2973T | A85V | 859 | 1 | 1500 | 0.38 |
| 14925 | Wave 3 | nsp12_2 | TRUE | C14925T | V486V | 128 | 0 | 849 | 0.21 |
| 16985 | Wave 3 | nsp13 | FALSE | C16985T | T250I | 98 | 1 | 141 | 0.04 |
| 20262 | Wave 3 | nsp15 | TRUE | A20262G | L214L | 60 | 1 | 522 | 0.13 |
| 20312 | Wave 3 | nsp15 | FALSE | C20312T | A231V | 859 | 0 | 148 | 0.04 |
| 21597 | Wave 3 | S | FALSE | C21597T | S12F | 874 | 1 | 633 | 0.16 |
| 25785 | Wave 3 | ORF3a | FALSE | G25785T\|A | W131C\|* | 202 | 8 | 1864 | 0.47 |
| 28308 | Wave 3 | N | FALSE | C28308G\|T | A12G\|V | 857 | 1 | 160 | 0.04 |
| 29144 | Wave 3 | N | TRUE | C29144T | L291L | 874 | 0 | 606 | 0.15 |
| 29585 | Wave 3 | ORF10 | FALSE | C29585T | P10S | 97 | 2 | 701 | 0.18 |
| 29711 | Wave 3 | Not in ORF | Not in ORF | G29711T | NA | 220 | 0 | 596 | 0.15 |
| 922 | Wave 4 | nsp2 | TRUE | G922A | L39L | 549 | 0 | 223 | 0.06 |
| 3431 | Wave 4 | nsp3 | FALSE | G3431T | V238L | 552 | 0 | 426 | 0.11 |
| 5653 | Wave 4 | nsp3 | TRUE | T5653C | Y978Y | 479 | 0 | 109 | 0.03 |
| 5812 | Wave 4 | nsp3 | TRUE | C5812T | D1031D | 117 | 1 | 903 | 0.23 |
| 5950 | Wave 4 | nsp3 | FALSE\|TRUE | G5950T\|A | K1077N\|K | 1110 | 12 | 1175 | 0.29 |
| 6255 | Wave 4 | nsp3 | FALSE | C6255T | A1179V | 552 | 0 | 543 | 0.14 |
| 7504 | Wave 4 | nsp3 | TRUE | C7504T | Y1595Y | 551 | 0 | 170 | 0.04 |
| 13176 | Wave 4 | nsp10 | FALSE | C13176T | T51I | 144 | 0 | 235 | 0.06 |
| 15720 | Wave 4 | nsp12_2 | TRUE | C15720T | D751D | 144 | 3 | 1377 | 0.35 |
| 18180 | Wave 4 | nsp14 | TRUE | G18180A | K47K | 144 | 0 | 33 | 0.01 |
| 18877 | Wave 4 | nsp14 | TRUE | C18877T | L280L | 617 | 1 | 23681 | 5.93 |
| 22418 | Wave 4 | S | FALSE | A22418G | T286A | 1 | 76 | 1 | 0 |
| 22444 | Wave 4 | S | TRUE | C22444T | D294D | 603 | 1 | 7893 | 1.98 |
| 23994 | Wave 4 | S | FALSE | A23994G | K811R | 1 | 93 | 9 | 0 |
| 24175 | Wave 4 | S | TRUE | T24175C | A871A | 536 | 0 | 59 | 0.01 |
| 25563 | Wave 4 | ORF3a | FALSE | G25563C\|T | Q57H | 631 | 10 | 89470 | 22.42 |
| 26735 | Wave 4 | M | TRUE | C26735T | Y71Y | 614 | 1 | 21029 | 5.27 |
| 28854 | Wave 4 | N | FALSE | C28854T | S194L | 611 | 3 | 23575 | 5.91 |

**Table S7.** Minor SNVs identified in Hong Kong cases (frequency in GISAID ≥1%).

| Position | Gene | Frequency in HK | (%) | Silent mutation | Frequency in GISAID | Frequency in GISAID (%) | Mutation (nucleotide) | Mutation (amino acid) |
| --- | --- | --- | --- | --- | --- | --- | --- | --- |
| 28883 | N | 25 | 1.56 | FALSE | 152044 | 38.09 | G28883C | G610R |
| 26801 | M | 10 | 0.62 | TRUE | 86175 | 21.59 | C26801G\|T | L279L |
| 22227 | S | 11 | 0.69 | FALSE | 85789 | 21.49 | C22227T | A665V |
| 21255 | nsp16 | 9 | 0.56 | TRUE | 85524 | 21.43 | G21255T\|C | A597A |
| 6286 | nsp3 | 12 | 0.75 | TRUE | 85407 | 21.4 | C6286T | T3567T |
| 29645 | ORF10 | 10 | 0.62 | FALSE | 85145 | 21.33 | G29645T | V88L |
| 28932 | N | 9 | 0.56 | FALSE | 85057 | 21.31 | C28932A\|T | A659D\|V |
| 445 | nsp1 | 10 | 0.62 | TRUE | 84963 | 21.29 | T445C | V180V |
| 1059 | nsp2 | 13 | 0.81 | FALSE | 59725 | 14.96 | C1059T | T254I |
| 27944 | ORF8 | 7 | 0.44 | TRUE | 57276 | 14.35 | C27944T | H51H |
| 23604 | S | 15 | 0.94 | FALSE | 56514 | 14.16 | C23604T\|G\|A | P2042L\|R\|H |
| 23063 | S | 16 | 1 | FALSE | 53756 | 13.47 | A23063T\|G | N1501Y\|D |
| 5986 | nsp3 | 13 | 0.81 | TRUE | 53506 | 13.41 | C5986T | F3267F |
| 28977 | N | 16 | 1 | FALSE | 53437 | 13.39 | C28977T | S704F |
| 3267 | nsp3 | 18 | 1.12 | FALSE | 52970 | 13.27 | C3267T | T548I |
| 14676 | nsp12_2 | 12 | 0.75 | TRUE | 52930 | 13.26 | C14676T | P1209P |
| 23709 | S | 11 | 0.69 | FALSE | 52646 | 13.19 | C23709T | T2147I |
| 27972 | ORF8 | 12 | 0.75 | FALSE | 52586 | 13.18 | C27972T | Q79* |
| 24914 | S | 12 | 0.75 | FALSE | 52551 | 13.17 | G24914T\|C | D3352Y\|H |
| 15279 | nsp12_2 | 12 | 0.75 | TRUE | 52500 | 13.15 | C15279T | H1812H |
| 23271 | S | 11 | 0.69 | FALSE | 52472 | 13.15 | C23271A | A1709D |
| 28048 | ORF8 | 12 | 0.75 | FALSE | 52430 | 13.14 | G28048A\|T | R155K\|I |
| 24506 | S | 11 | 0.69 | FALSE | 52370 | 13.12 | T24506G | S2944A |
| 16176 | nsp12_2 | 12 | 0.75 | TRUE | 52366 | 13.12 | T16176C | T2709T |
| 28111 | ORF8 | 10 | 0.62 | FALSE | 52347 | 13.12 | A28111G | Y218C |
| 6954 | nsp3 | 11 | 0.69 | FALSE | 52344 | 13.11 | T6954C | I4235T |
| 5388 | nsp3 | 7 | 0.44 | FALSE | 52288 | 13.1 | C5388A | A2669D |
| 913 | nsp2 | 14 | 0.87 | TRUE | 52196 | 13.08 | C913T | S108S |
| 204 | Not in ORF | 5 | 0.31 | Not in ORF | 47971 | 12.02 | G204T\|A | NA |
| 21614 | S | 4 | 0.25 | FALSE | 39312 | 9.85 | C21614T | L52F |
| 20268 | nsp15 | 7 | 0.44 | TRUE | 26684 | 6.69 | A20268G | L648L |
| 27964 | ORF8 | 6 | 0.37 | FALSE | 24942 | 6.25 | C27964T | S71L |
| 28869 | N | 7 | 0.44 | FALSE | 23983 | 6.01 | C28869T | P596L |
| 313 | nsp1 | 24 | 1.5 | TRUE | 21829 | 5.47 | C313T | L48L |
| 22992 | S | 9 | 0.56 | FALSE | 21826 | 5.47 | G22992C\|A\|T | S1430T\|N\|I |
| 10319 | nsp5 | 6 | 0.37 | FALSE | 20462 | 5.13 | C10319T\|A | L265F\|I |
| 11083 | nsp6 | 38 | 2.37 | FALSE\|TRUE | 20168 | 5.05 | G11083T\|A | L111F\|L |
| 17615 | nsp13 | 9 | 0.56 | FALSE | 19741 | 4.95 | A17615G | K1379R |
| 28975 | N | 5 | 0.31 | FALSE | 19390 | 4.86 | G28975C\|T\|A | M702I |
| 21304 | nsp16 | 6 | 0.37 | FALSE | 18702 | 4.69 | C21304T | R646C |
| 18424 | nsp14 | 4 | 0.25 | FALSE | 17916 | 4.49 | A18424G | N385D |
| 25907 | ORF3a | 4 | 0.25 | FALSE | 17673 | 4.43 | G25907T | G515V |
| 28472 | N | 4 | 0.25 | FALSE | 17473 | 4.38 | C28472T | P199S |
| 14805 | nsp12_2 | 4 | 0.25 | TRUE | 14592 | 3.66 | C14805T | Y1338Y |
| 4543 | nsp3 | 5 | 0.31 | TRUE | 11630 | 2.91 | C4543T | T1824T |
| 25710 | ORF3a | 3 | 0.19 | TRUE | 11496 | 2.88 | C25710T | L318L |
| 15766 | nsp12_2 | 2 | 0.12 | FALSE | 11473 | 2.87 | G15766T | V2299L |
| 17019 | nsp13 | 5 | 0.31 | FALSE\|TRUE | 11208 | 2.81 | G17019T\|A | E783D\|E |
| 9526 | nsp4 | 4 | 0.25 | FALSE | 11146 | 2.79 | G9526T\|C | M972I |
| 13993 | nsp12_2 | 2 | 0.12 | FALSE | 11064 | 2.77 | G13993T | A526S |
| 11497 | nsp6 | 2 | 0.12 | TRUE | 11042 | 2.77 | C11497T | Y525Y |
| 26876 | M | 2 | 0.12 | TRUE | 10983 | 2.75 | T26876C | I354I |
| 16889 | nsp13 | 2 | 0.12 | FALSE | 10934 | 2.74 | A16889G | K653R |
| 29399 | N | 2 | 0.12 | FALSE | 10916 | 2.73 | G29399A | A1126T |
| 23401 | S | 3 | 0.19 | FALSE | 10620 | 2.66 | G23401T | Q1839H |
| 5629 | nsp3 | 2 | 0.12 | TRUE | 10398 | 2.61 | G5629T | T2910T |
| 15324 | nsp12_2 | 4 | 0.25 | TRUE | 10168 | 2.55 | C15324T | N1857N |
| 29734 | Not in ORF | 5 | 0.31 | Not in ORF | 9939 | 2.49 | G29734T\|C | NA |
| 222 | Not in ORF | 2 | 0.12 | Not in ORF | 9834 | 2.46 | C222T | NA |
| 17104 | nsp13 | 5 | 0.31 | FALSE | 8996 | 2.25 | C17104T | H868Y |
| 22879 | S | 3 | 0.19 | FALSE | 8965 | 2.25 | C22879A | N1317K |
| 29366 | N | 3 | 0.19 | FALSE | 8923 | 2.24 | C29366T | P1093S |
| 7767 | nsp3 | 12 | 0.75 | FALSE | 8618 | 2.16 | T7767C | I5048T |
| 8047 | nsp3 | 3 | 0.19 | TRUE | 8583 | 2.15 | C8047T | Y5328Y |
| 8083 | nsp3 | 5 | 0.31 | FALSE | 8156 | 2.04 | G8083A | M5364I |
| 20661 | nsp16 | 3 | 0.19 | TRUE | 8129 | 2.04 | T20661C | S3S |
| 29402 | N | 10 | 0.62 | FALSE | 7511 | 1.88 | G29402T\|C | D1129Y\|H |
| 27800 | ORF7b | 3 | 0.19 | TRUE | 7440 | 1.86 | C27800A | A39A |
| 28725 | N | 3 | 0.19 | FALSE | 7311 | 1.83 | C28725T | P452L |
| 9286 | nsp4 | 17 | 1.06 | TRUE | 7271 | 1.82 | C9286T | N732N |
| 10097 | nsp5 | 6 | 0.37 | FALSE | 6822 | 1.71 | G10097A\|T\|C | G43S\|C\|R |
| 18028 | nsp13 | 6 | 0.37 | FALSE | 6674 | 1.67 | G18028T | A1792S |
| 21855 | S | 5 | 0.31 | FALSE | 6653 | 1.67 | C21855T | S293F |
| 21575 | S | 25 | 1.56 | FALSE | 6592 | 1.65 | C21575T | L13F |
| 12988 | nsp9 | 5 | 0.31 | FALSE | 6563 | 1.64 | G12988T\|C | M303I |
| 26972 | M | 3 | 0.19 | TRUE | 6559 | 1.64 | T26972C | R450R |
| 15598 | nsp12_2 | 3 | 0.19 | FALSE | 6554 | 1.64 | G15598A | V2131I |
| 24910 | S | 5 | 0.31 | TRUE | 6544 | 1.64 | T24910C\|G | T3348T |
| 2453 | nsp2 | 4 | 0.25 | FALSE | 6270 | 1.57 | C2453T | L1648F |
| 28651 | N | 4 | 0.25 | TRUE | 6143 | 1.54 | C28651T | N378N |
| 28887 | N | 8 | 0.5 | FALSE | 6124 | 1.53 | C28887T | T614I |
| 19839 | nsp15 | 2 | 0.12 | TRUE | 6093 | 1.53 | T19839C | N219N |
| 23731 | S | 4 | 0.25 | TRUE | 6068 | 1.52 | C23731T | T2169T |
| 10323 | nsp5 | 11 | 0.69 | FALSE | 5911 | 1.48 | A10323G | K269R |
| 11396 | nsp6 | 3 | 0.19 | FALSE | 5509 | 1.38 | C11396T | L424F |
| 2416 | nsp2 | 6 | 0.37 | TRUE | 5362 | 1.34 | C2416T | Y1611Y |
| 10870 | nsp5 | 5 | 0.31 | TRUE | 5156 | 1.29 | G10870T\|A | L816L |
| 9745 | nsp4 | 3 | 0.19 | TRUE | 5069 | 1.27 | C9745T | Y1191Y |
| 20451 | nsp15 | 4 | 0.25 | TRUE | 4978 | 1.25 | C20451T | N831N |
| 22346 | S | 4 | 0.25 | FALSE | 4665 | 1.17 | G22346T | A784S |
| 28087 | ORF8 | 2 | 0.12 | FALSE | 4662 | 1.17 | C28087T | A194V |
| 26424 | E | 7 | 0.44 | TRUE | 4656 | 1.17 | T26424C | S180S |
| 8603 | nsp4 | 3 | 0.19 | FALSE | 4626 | 1.16 | T8603C | F49L |
| 13536 | nsp12_2 | 3 | 0.19 | TRUE | 4547 | 1.14 | C13536T | Y69Y |
| 15480 | nsp12_2 | 3 | 0.19 | TRUE | 4530 | 1.13 | C15480A\|T | T2013T |
| 3177 | nsp3 | 3 | 0.19 | FALSE | 4398 | 1.1 | C3177T | P458L |
| 8917 | nsp4 | 8 | 0.5 | TRUE | 4375 | 1.1 | C8917T | F363F |
| 4002 | nsp3 | 4 | 0.25 | FALSE | 4288 | 1.07 | C4002T | T1283I |
| 19524 | nsp14 | 3 | 0.19 | TRUE | 4175 | 1.05 | C19524T | L1485L |
| 29179 | N | 5 | 0.31 | TRUE | 4162 | 1.04 | G29179T\|A\|C | P906P |
| 25437 | ORF3a | 4 | 0.25 | FALSE | 4088 | 1.02 | G25437T | L45F |
| 22388 | S | 2 | 0.12 | TRUE | 4038 | 1.01 | C22388T | L826L |
| 28253 | ORF8 | 43 | 2.69 | TRUE | 3982 | 1 | C28253T | F360F |

**Table S8.** Estimation on bottleneck size of transmission pairs.

| **Transmission pair** | **Variant calling threshold** | **Donor** | **Recipient** | **Bottleneck size** | **CI lower** | **CI upper** |
| --- | --- | --- | --- | --- | --- | --- |
| Cluster_fam_1122 | 0.03 | 8773 | 8772 | 3 | 1 | 10 |
| Cluster_fam_1166 | 0.03 | 9042 | 9041 | 2 | 1 | 4 |
| Cluster_fam_197 | 0.03 | 1905 | 2168 | 1 | 0 | 9 |
| Cluster_fam_222 | 0.03 | 2172 | 2317 | 1 | 0 | 27 |
| Cluster_fam_293 | 0.03 | 2735 | 2609 | 1 | 0 | 202 |
| Cluster_fam_336 | 0.03 | 2989 | 2962 | 1 | 0 | 13 |
| Cluster_fam_509 | 0.03 | 3970 | 3612 | 1 | 0 | 22 |
| Cluster_fam_562 | 0.03 | 4306 | 4307 | NA | NA | NA |
| Cluster_fam_718 | 0.03 | 5399 | 5444 | 1 | 0 | 13 |
| Cluster_fam_730 | 0.03 | 5539 | 5577 | NA | NA | NA |
| Cluster_friends_25 | 0.03 | 1839 | 2047 | 1 | 0 | 10 |
| Cluster_roommate_08 | 0.03 | 2721 | 2545 | 1 | 0 | 5 |
| Cluster_roommate_21 | 0.03 | 4075 | 4208 | 1 | 0 | 4 |

**Table S9.**
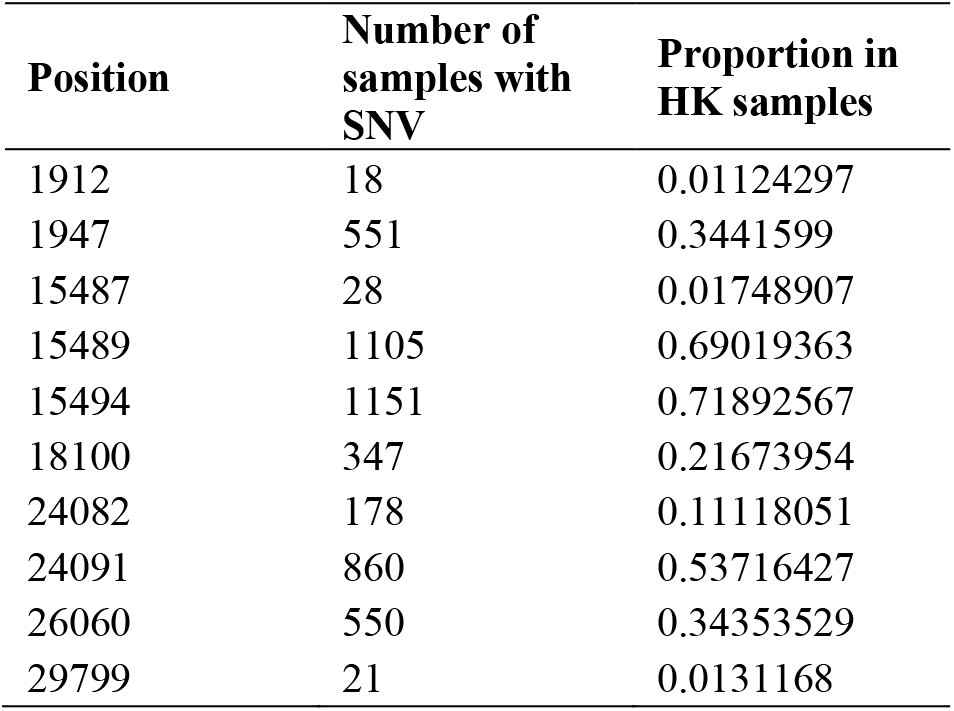
Highly shared variant sites (allele frequency ≥3% and were found in >1% of the HK samples) located within or related to PCR primer binding regions.

**Table S10.** Gene annotation of SARS-CoV-2 Genome (nucleotide positions base on reference sequence Wuhan-Hu-1, GenBank: MN908947.3).

| **Gene segment** | **Start** | **Stop** |
| --- | --- | --- |
| nsp1 | 266 | 805 |
| nsp2 | 806 | 2719 |
| nsp3 | 2720 | 8554 |
| nsp4 | 8555 | 10054 |
| nsp5 | 10055 | 10972 |
| nsp6 | 10973 | 11842 |
| nsp7 | 11843 | 12091 |
| nsp8 | 12092 | 12685 |
| nsp9 | 12686 | 13024 |
| nsp10 | 13025 | 13441 |
| nsp12_1 | 13442 | 13468 |
| nsp12_2 | 13468 | 16236 |
| nsp13 | 16237 | 18039 |
| nsp14 | 18040 | 19620 |
| nsp15 | 19621 | 20658 |
| nsp16 | 20659 | 21555 |
| S | 21563 | 25384 |
| ORF3a | 25393 | 26220 |
| E | 26245 | 26472 |
| M | 26523 | 27191 |
| ORF6 | 27202 | 27387 |
| ORF7a | 27394 | 27753 |
| ORF7b | 27762 | 27887 |
| ORF8 | 27894 | 28259 |
| N | 28274 | 29533 |
| ORF10 | 29558 | 29674 |

**Data S1. (Data S1.csv)**

Origins of imported cases in Hong Kong.

**Data S2. (Data S2.csv)**

Sample list with waves, NextClade and PANGO lineage designations.

**Data S3. (Data S3.csv)**

Summary of Hong Kong monophyletic clades.

**Data S4. (Data S4.pdf)**

Acknowledgements to sequences obtained from GISAID (accessed on 11-June-2021).

